# Longitudinal Participation and Cognitive Score Interpretation in the J-TRC Webstudy

**DOI:** 10.64898/2026.11.06.26367280

**Authors:** Kenichiro Sato, Yoshiki Niimi, Ryoko Ihara, Kazushi Suzuki, Atsushi Iwata, Takeshi Iwatsubo

## Abstract

**INTRODUCTION:** Web-based registries support remote cognitive assessment for Alzheimer’s disease research, but participant-driven reassessment raises questions about long-term data yield and interpretation of repeated scores. We evaluated data contribution, selective observation, and retest-dependent estimates of cognitive change in the Japanese Trial-Ready Cohort (J-TRC) Webstudy.

**METHODS:** We analyzed over six years of J-TRC Webstudy data (launched October 2019; registrations through March 31, 2025; assessments through May 31, 2026). We examined data contribution, inactivity, reactivation, score-associated observation, first-exposure contrasts, and sensitivity to retest models and estimators.

**RESULTS:** Among 14,912 registered accounts, 10,863 participants were eligible; 10,645 (98.0%) completed the Cognitive Function Instrument (CFI), 6,419 (59.1%) completed Cogstate, and 5,792 (53.3%) contributed assessments on multiple days. Among those with sufficient opportunity, 19.4% and 14.1% contributed data at or beyond three and five years. After 365 days without assessment, the estimated probability of resuming within two years was 7.2%. Worse prior cognitive scores were associated with lower odds of subsequent assessment (adjusted odds ratios per standard deviation: CFI, 0.952; Cogstate, 0.845). By assessment 12, returnee changes were +0.43 and +1.14 favorable baseline standard deviations (CFI, Cogstate), versus −0.06 and +0.08 in first-exposure contrasts. Marginal CFI time estimates differed under categorical versus flexible retest adjustment (+0.080 versus −0.227 standard deviations/year).

**DISCUSSION:** High initial completion was accompanied by limited long-term data contribution. Subsequent assessment was associated with prior scores, and estimated time trends varied by retest model. Reporting assessment histories and testing model sensitivity are important for interpreting repeated online cognitive scores.

## Introduction

Identifying individuals at risk of Alzheimer’s disease (AD) and maintaining their participation over extended periods are important challenges for observational research and prevention trials [Grill2010(PMID: 21172069); Aisen2020(PMID: 32920621)]. Web-based registries offer opportunities to recruit large numbers of participants, collect cognitive and functional assessments remotely, and facilitate referral to in-person studies, potentially reducing geographical and logistical barriers [Weiner2018(PMID: 29754989); Walter2020(PMID: 32920623)]. Several web-based research initiatives, including the Brain Health Registry (BHR) [Weiner2018(PMID: 29754989)], Alzheimer Prevention Trials (APT) Webstudy [Walter2020(PMID: 32920623)], Healthy Brain Project (HBP) [Lim2019(PMID: 30909237)], and PROTECT study [Wesnes2017(PMID: 28128869)], have demonstrated the feasibility of large-scale recruitment and remote cognitive assessment.

The Japanese Trial-Ready Cohort (J-TRC) Webstudy, launched in October 2019 as the web-based component of the J-TRC program and adapted from the APT Webstudy, was established to identify and longitudinally monitor potential participants for AD prevention trials in Japan [Iwatsubo2024(PMID: 38842037); Sato2021(PMID: 34292159)]. The J-TRC Webstudy combines online registration with repeated self-reported cognitive-functional assessments using the Cognitive Function Instrument (CFI) and optional computerized testing using the Cogstate Brief Battery [Sato2021(PMID: 33778148)]. The J-TRC program has used amyloid-risk estimates derived from models trained on A4 screening data and applied to Webstudy participants’ demographic characteristics and available CFI and Cogstate scores to prioritize invitations to the onsite study [Sato2021(PMID: 33778148); Nakashima2026(PMID: 41536849)]. Thus, the interpretation of online cognitive assessments is relevant to both longitudinal monitoring and risk stratification within a trial-ready cohort.

Beyond initial recruitment, the longitudinal value of trial-ready registries depends on sustained data contribution and the interpretation of repeated assessments. Sustaining participation over time remains challenging: the Brain Health Registry reported follow-up return rates of 41% at six months and 25% at 24 months [Weiner2018(PMID: 29754989)]. Subsequent research has examined participant characteristics associated with repeated assessment completion, including sociodemographic differences in online registry participation [Ashford2020(PMID: 32478165); Ashford2024(PMID: 38188606)]. However, return rates alone do not characterize irregular assessment intervals, prolonged inactivity followed by reactivation, or how these assessment histories relate to the scores that are observed. Evaluating participation and score interpretation together is therefore important when reassessment is participant-driven.

Repeated assessments pose an additional challenge for interpreting cognitive and cognitive-functional scores. Repeated exposure can improve cognitive test performance [Stricker2020(PMID: 32010922); Murchland2026(PMID: 42224199)], and estimates of longitudinal cognitive change can vary depending on how retest exposure is modeled [Hoffman2011(PMID: 21639642); Vivot2016(PMID: 26825924); Chen2023(PMID: 37199336)]. At the same time, reduced practice effects have been investigated as potential markers of cognitive impairment and future decline [Jutten2020(PMID: 32671181)]. Changes in self-reported scores may also reflect changes in reporting behavior [Shrout2018(PMID: 29255039)]. In addition, participants with poorer cognitive performance may be less likely to complete subsequent assessments [Hernandez2024(PMID: 38453191)], potentially altering the composition of the observed sample. Together, retest effects and selective follow-up complicate the interpretation of observed score trajectories [Vivot2016(PMID: 26825924); Gasparini2020(PMID: 31894164); Weuve2015(PMID: 26397878)].

Using over six years of J-TRC Webstudy data, we evaluated the yield and interpretation of longitudinal cognitive data collected through participant-driven reassessment. We first characterized progression from registration to repeated assessment, multiyear data contribution, inactivity, and reactivation. We then examined how prior cognitive scores and assessment histories were associated with subsequent observation. Finally, we compared score changes among returnees with contemporaneous first-exposure contrasts and examined the sensitivity of estimated time trends to retest specifications and statistical estimators.

## Methods

### Study design and participants

We conducted a longitudinal observational study of the ongoing Japanese Trial-Ready Cohort (J-TRC) Webstudy, launched in October 2019 for AD prevention research [Sato2021(PMID: 34292159); Sato2021(PMID: 33778148)]. The Webstudy’s public landing page is shown in Figure 1A. Participants register and consent online; the Cognitive Function Instrument (CFI) is integrated into the assessment process, whereas Cogstate testing is optional. The same instrument becomes available for voluntary reassessment 90 days after completion, without scheduled study visits (Figure 1B).

**Figure 1.**
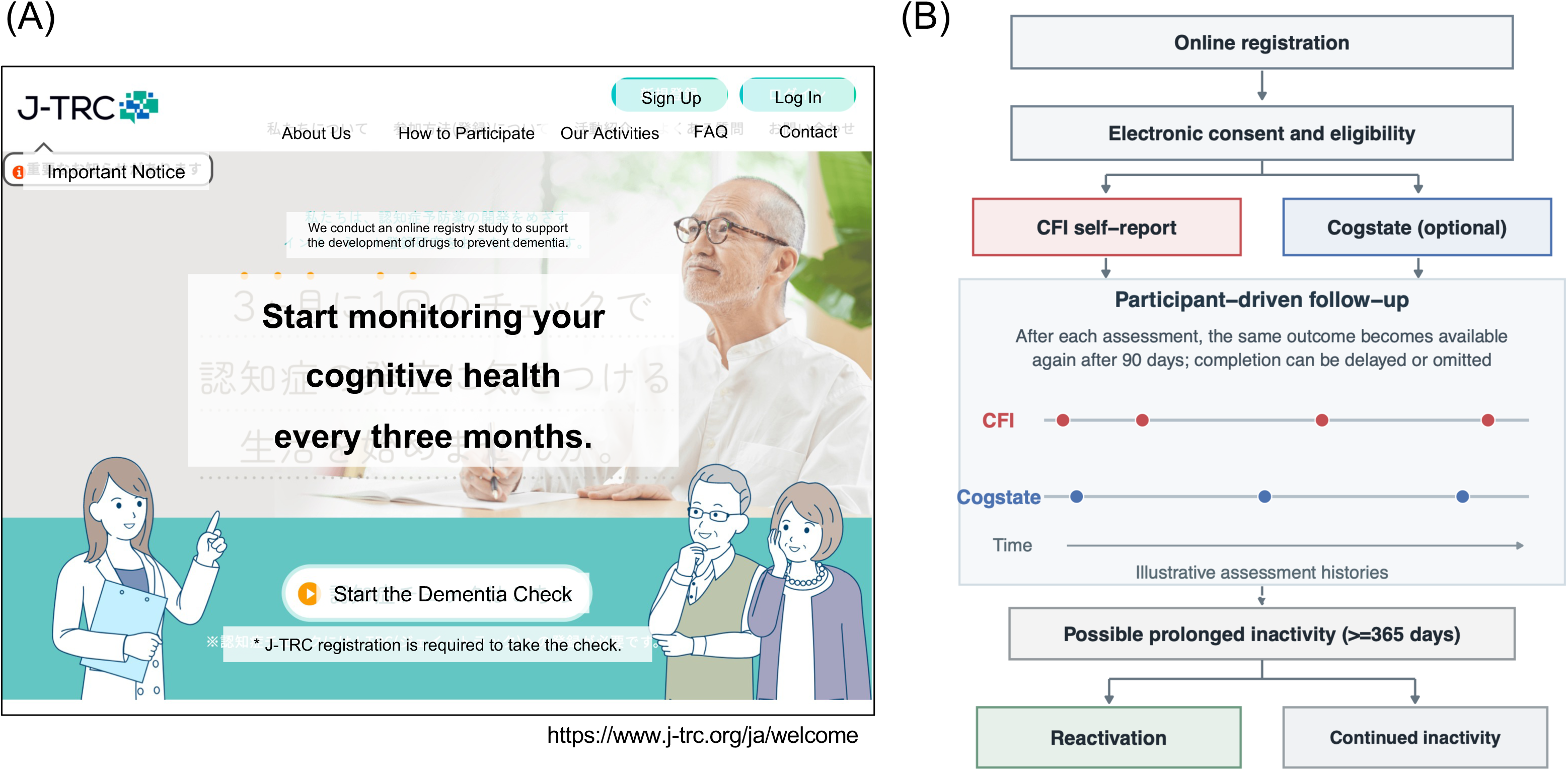
J-TRC Webstudy interface and participant-driven assessment process. (A) Representative screenshot of the Japanese-language J-TRC Webstudy public landing page (https://www.j-trc.org/ja/welcome), captured in October 2026. The screenshot illustrates the public-facing interface and was obtained after the administrative observation cutoff of the present study. English translations are overlaid on the original Japanese text for presentation purposes. The original website is in Japanese. (B) Schematic overview of online registration, electronic consent and eligibility assessment, CFI self-report, optional Cogstate assessment, and participant-driven follow-up. Following completion of an assessment, reassessment using the same instrument becomes available after 90 days; subsequent completion is voluntary and does not follow a fixed visit schedule. Illustrative assessment histories demonstrate irregular intervals, possible prolonged inactivity (at least 365 days without an assessment), and subsequent reactivation. These pathways are conceptual and do not represent individual participant data. **Abbreviations**: CFI, Cognitive Function Instrument; J-TRC, Japanese Trial-Ready Cohort.

For Cogstate, participants received an email notification approximately three months after completing an assessment, when retesting became available. This notification was sent once per completed assessment and was not repeated if the participant did not retest.

We included registrations through March 31, 2025, allowing at least 12 months of potential follow-up, and assessments through May 31, 2026. The University of Tokyo Hospital Institutional Review Board approved the Webstudy (2019132NI). All participants included in the eligible analytical cohort provided electronic informed consent.

Initial recruitment activities included a press release at the Webstudy launch in October 2019, followed by newspaper and online media coverage. Additional recruitment efforts included search-engine listing and banner advertisements, paid newspaper advertisements, and email invitations to participants in an existing cognitive registry. The timing and short-term recruitment effects of these activities have been reported previously [Sato2021(PMID: 34292159)]. A further public announcement highlighting progress in the J-TRC program and encouraging participation in the Webstudy was issued in February 2022 [UTokyoAMED2022].

We distinguished registered accounts, accounts with explicit affirmative consent, participants with ascertainable eligibility information, and eligible consented participants. The primary registry-level analytical population comprised individuals who had provided affirmative consent, had ascertainable age and self-reported AD status, were aged 50–85 years at registration, and did not report a previous AD diagnosis. Completion of all registration workflow stages or a valid cognitive assessment was not required for inclusion in this population. Assessment-specific populations were subsequently defined according to the availability of valid CFI or Cogstate records, and regression analyses were restricted to participants with the variables required by each model. Accordingly, the denominators for registered accounts, eligible participants, participants completing valid assessments, and longitudinal modeling populations were distinguished throughout the analyses (Figure 2B; Table 1; Supplementary Table S1).

**Figure 2.**
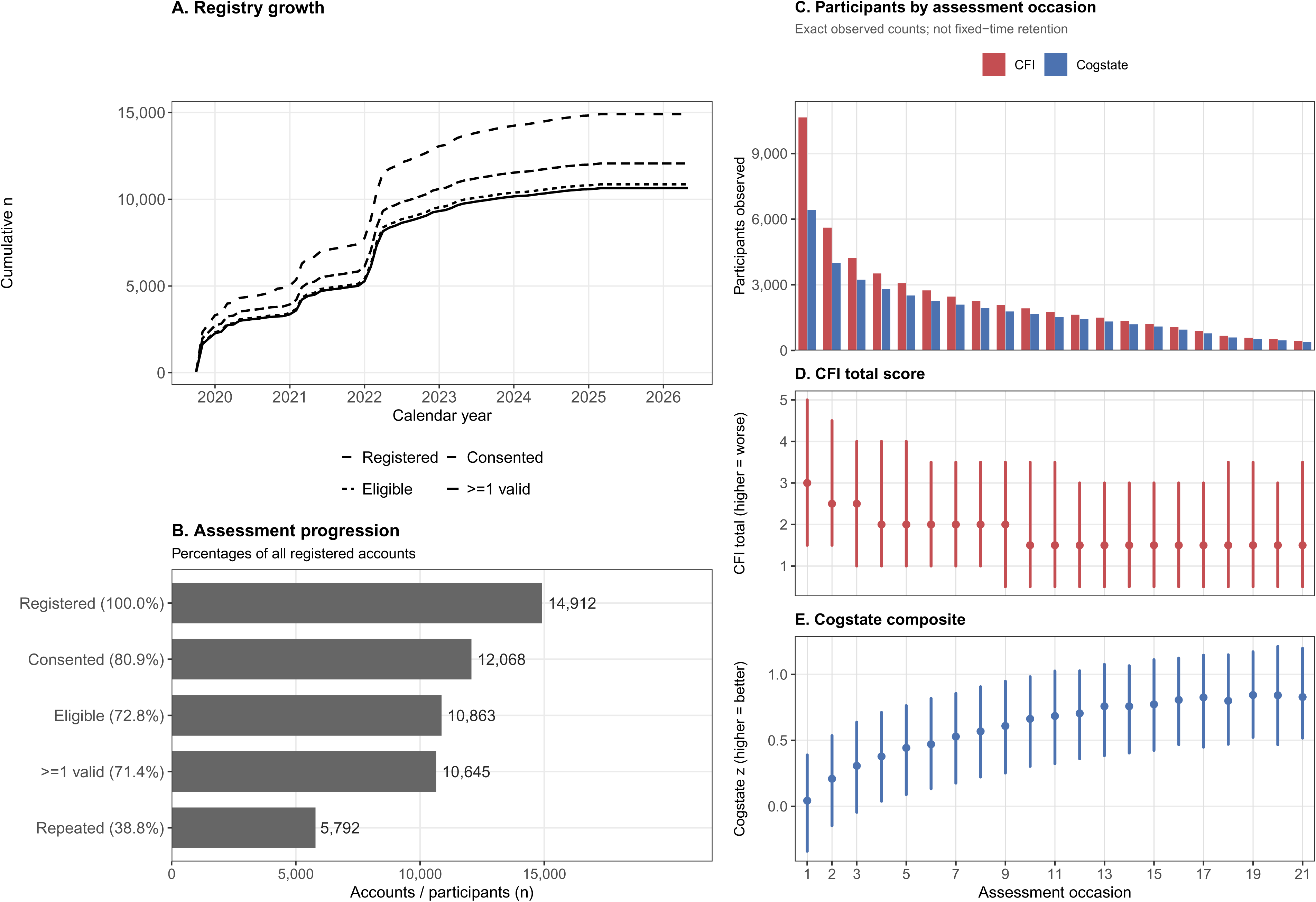
Registry growth, assessment progression, and repeated-score distributions. (A) Cumulative numbers of registered accounts, accounts with explicit affirmative consent, eligible consented participants, and participants completing at least one valid assessment, by calendar date. (B) Registry-to-assessment progression showing registered accounts, explicit affirmative consent, eligibility, completion of at least one valid assessment, and repeated assessment activity on at least two distinct calendar days. Percentages in panel B use all registered accounts (n = 14,912) as the denominator. Corresponding percentages based on the eligible consented cohort are provided in Table 1. (C) Numbers of participants completing each assessment occasion, from the first through the 21st, separately for CFI and Cogstate. (D) Observed CFI total scores by assessment occasion. (E) Observed Cogstate composite scores by assessment occasion. In panels D and E, filled circles represent medians and vertical intervals represent interquartile ranges. Assessment occasion denotes the cumulative number of valid assessments completed by an individual participant for the respective instrument, rather than a scheduled follow-up visit or fixed elapsed time. Higher CFI scores indicate greater self-reported cognitive-functional difficulties; higher Cogstate composite scores indicate favorable performance under the analytic scoring convention. Distributions at each occasion reflect participants completing that assessment; within-person changes among returnees are shown in Figure 4A. Registrations were restricted to March 31, 2025, and assessment data were available through May 31, 2026. **Abbreviations**: CFI, Cognitive Function Instrument; IQR, interquartile range; J-TRC, Japanese Trial-Ready Cohort; n, number.

### Cognitive assessments and scoring

The CFI is a 15-item self-reported measure of cognitive and functional difficulties, with higher scores indicating greater reported difficulties [Amariglio2015(PMID: 25706191); Walter2020(PMID: 32920625)]. Item responses were coded as 0 for “No,” 0.5 for “Maybe,” and 1 for “Yes” [Amariglio2015(PMID: 25706191)]. A “Does not apply” response counted toward questionnaire completion but did not contribute a numerical item score. A valid CFI record required completion-valid responses to all 15 items, an assessment date, and an available recorded total. The primary CFI score was reconstructed as the sum of scorable item responses without prorating. Two separate sensitivity analyses were conducted: one restricted to assessments with all 15 items numerically scorable, and another using total scores prorated to 15 items according to the number of scorable responses. Exact duplicate CFI records were removed, and when multiple valid assessments occurred on the same calendar day, the latest record was retained.

Cogstate comprised Detection (DET; psychomotor speed), Identification (IDN; visual attention), One Card Learning (OCL; visual recognition learning), and One Back (ONB; working memory) [Maruff2013(PMID: 25566378); Sato2021(PMID: 33778148)]. Standard primary measures are reaction time for DET and IDN and accuracy for OCL and ONB. Four task scores were exported, treated as higher-is-better without further sign reversal, and re-standardized using the task-specific means and standard deviations at participants’ first observed Cogstate assessments. Their average defined the study-specific composite when at least two task scores were available. Duplicate sessions were removed. For longitudinal analyses, the Cogstate composite and CFI were scaled by their respective first-assessment standard deviations, with CFI changes sign-reversed so that positive values indicated improvement.

### Participation and longitudinal data contribution

We quantified progression from registration to consent, eligibility, initial assessment, repeated assessment, and long-term data contribution. Assessment activity was defined using calendar dates; CFI and Cogstate assessments occurring on the same day contributed one day to the combined activity count. Repeated participation was defined as activity on at least two distinct calendar days. Longitudinal data contribution was assessed by determining whether participants provided at least one assessment at or beyond 12, 36, and 60 months after registration. For each threshold, the denominator was restricted to eligible participants whose registration dates allowed sufficient observation opportunity before May 31, 2026. We also calculated the proportion with assessment activity during the final 365 days of observation. These measures describe observed data contribution rather than adherence to scheduled visits, which did not exist in the Webstudy.

Assessment occasion was each participant’s cumulative number of valid assessments for an instrument, not a scheduled visit. We summarized occasion-specific participant counts and score medians and interquartile ranges (Figure 2C–E) and examined their relationship with elapsed time (Supplementary Figure S1). Participation was compared between 2019–2021 and 2022–2023 registration cohorts within a common 24-month window. Characteristics across participation stages were described using standardized mean differences (Supplementary Tables S1 and S2), without seeking independent determinants of engagement.

### Dynamic participation and reactivation

To characterize changing participation states, we divided each participant’s follow-up period into nonoverlapping 90-day intervals starting from their registration date. At the end of each complete interval, eligible participants were classified into three mutually exclusive states: no previous valid assessment, recently active (an assessment less than 365 days earlier), or inactive (previous assessment activity followed by at least 365 consecutive days without assessment). State proportions were calculated among participants with sufficient observation opportunity at each interval endpoint. Incomplete terminal intervals were excluded.

Kaplan–Meier analyses estimated time from the first assessment to the first 365-day gap without an assessment, with administrative censoring. Reactivation was defined as the next assessment after such a gap; time zero was when the first qualifying gap reached 365 days. Cumulative reactivation probabilities used Kaplan–Meier estimation with log-log confidence intervals (Figure 3A,B). For reactivation episodes with at least 365 days of subsequent observation opportunity, we assessed whether assessments occurred on at least two additional days within the following year. Sensitivity analyses varied the inactivity threshold (180 or 730 days) and registration-anchored panel width (60 or 120 days; Supplementary Figure S2).

**Figure 3.**
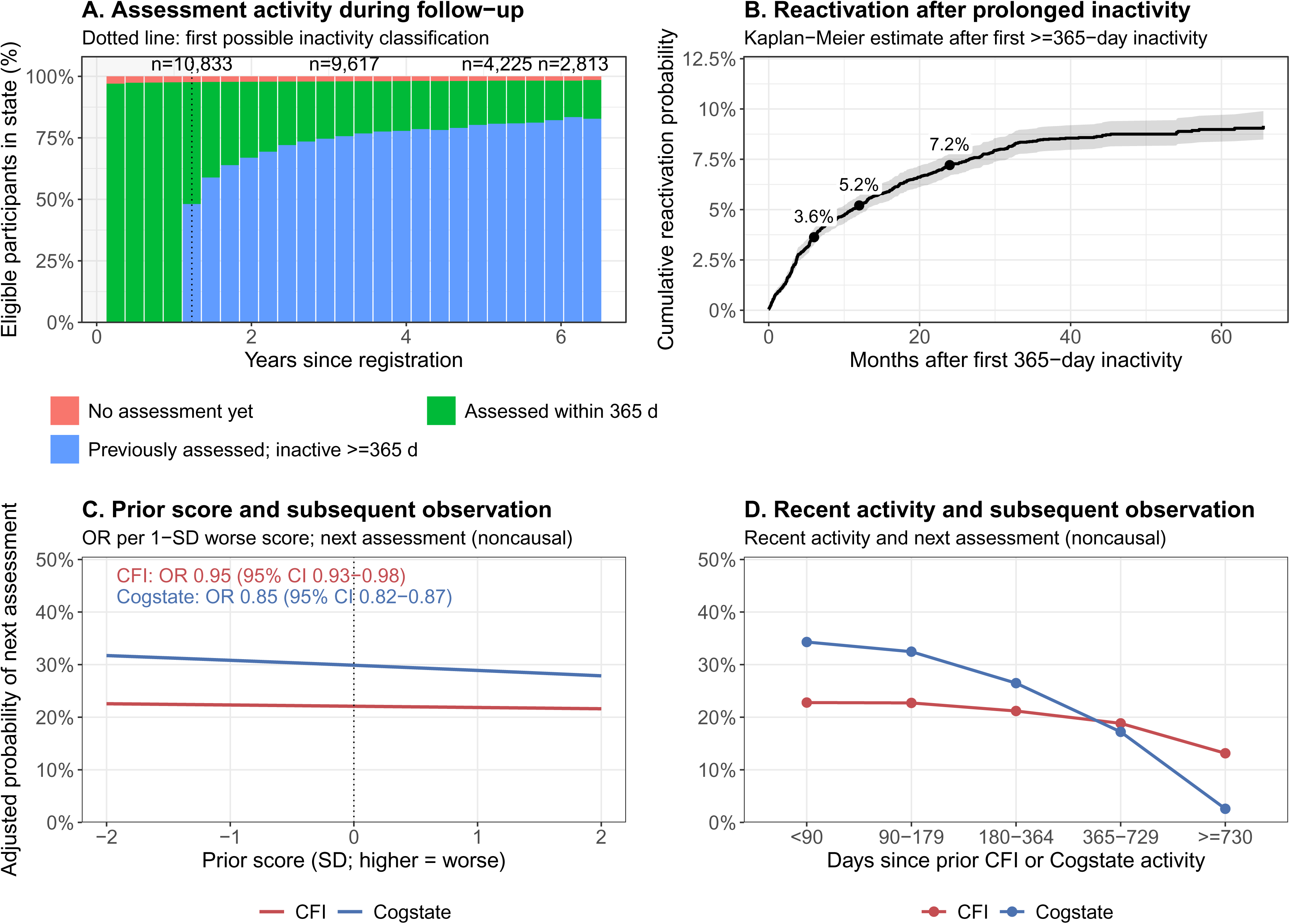
Dynamic participation, reactivation, and associations with subsequent assessment observation. **(A)** Distribution of participation states at registration-anchored 90-day endpoints among eligible participants with sufficient observation opportunity. States comprise no previous assessment, assessment within the preceding 365 days, and previous assessment followed by at least 365 days of inactivity. The vertical dotted line indicates the first time at which the prolonged-inactivity criterion could be met. Labels above selected intervals indicate the numbers of eligible participants contributing to the respective denominators. **(B)** Kaplan–Meier cumulative probability of reactivation after the first episode of at least 365 consecutive days without a valid assessment. Time is measured from attainment of the inactivity threshold until subsequent assessment or administrative censoring. The shaded region represents the 95% confidence interval. Cumulative reactivation probabilities at approximately 6, 12, and 24 months were 3.6%, 5.2%, and 7.2%, with corresponding numbers at risk of 8,456, 7,983, and 6,896. These risk-set counts are not simple denominators for the cumulative probabilities. **(C)** Model-standardized probability of subsequent assessment observation according to the previously observed score, expressed in standard-deviation units with higher values indicating worse performance for both instruments. Annotated odds ratios represent the adjusted association per 1-SD worse prior score, with participant-clustered 95% confidence intervals. **(D)** Model-standardized probability of subsequent assessment observation according to time since the most recent Webstudy assessment activity, irrespective of instrument. Predictions in panels C and D were averaged over the empirical covariate distribution of the respective analytical samples. **Abbreviations:** CFI, Cognitive Function Instrument; CI, confidence interval; OR, odds ratio; SD, standard deviation.

### Associations with subsequent assessment completion

We investigated whether previously observed scores and assessment histories were associated with the probability of subsequent assessment completion. For each instrument, outcome-specific rolling 90-day risk intervals were constructed beginning when reassessment became available, 90 days after the preceding assessment. The initial assessment was conditioned on, and subsequent intervals were classified according to whether an analyzable assessment was observed. We fitted logistic regression models for interval-specific observation, including elapsed time and its squared term, age, sex, educational attainment, registration year, the last observed score, prior assessment count, previous observation history, and days since the most recent Webstudy activity. Missing prior-score history was represented using a median-imputed value and a missingness indicator. Confidence intervals for the association between prior score and subsequent observation were obtained using participant-clustered robust sandwich estimators and expressed as odds ratios per 1-standard-deviation worse prior score. Model-standardized probabilities were obtained by averaging predictions over the empirical covariate distribution while varying the prior score or time since the most recent activity. Recency categories were used to summarize these standardized probabilities for presentation (Figure 3C,D).

### Analysis of longitudinal cognitive score patterns

The score analyses addressed three related but distinct quantities: within-person score change among observed returnees, contrasts between matched first-exposure assessments from different participants, and model-specific elapsed-time associations conditional on the specified retest terms. We compared retest-model specifications (Supplementary Figure S3; Supplementary Table S3), constructed contemporaneous first-exposure references, and examined sensitivity to selective observation. We examined these quantities separately because they capture different aspects of the observed score data. To contextualize score changes among repeatedly assessed participants, we constructed an internal reference using contemporaneous first-observed assessments from other participants (Figure 4A). This comparison is conceptually related to replacement-participant approaches [Elman2018(PMID: 30003138)], and provided an empirical benchmark for score changes observed among returnees. For returnee assessment occasions 2–12 occurring within 36 months of the participant’s first assessment, we calculated within-person score changes. Two distinct participants completing their first observed J-TRC assessment were separately matched to each returnee’s initial and subsequent assessment dates. Matching used greedy nearest-neighbor matching without replacement within each matching analysis, requiring the same sex, calendar dates within ±180 days, attained ages within ±2 years, and educational attainment within ±2 years. The reference contrast was calculated as the difference between the two independently matched first-exposure scores, whereas the returnee change was calculated within the same participant.

**Figure 4.**
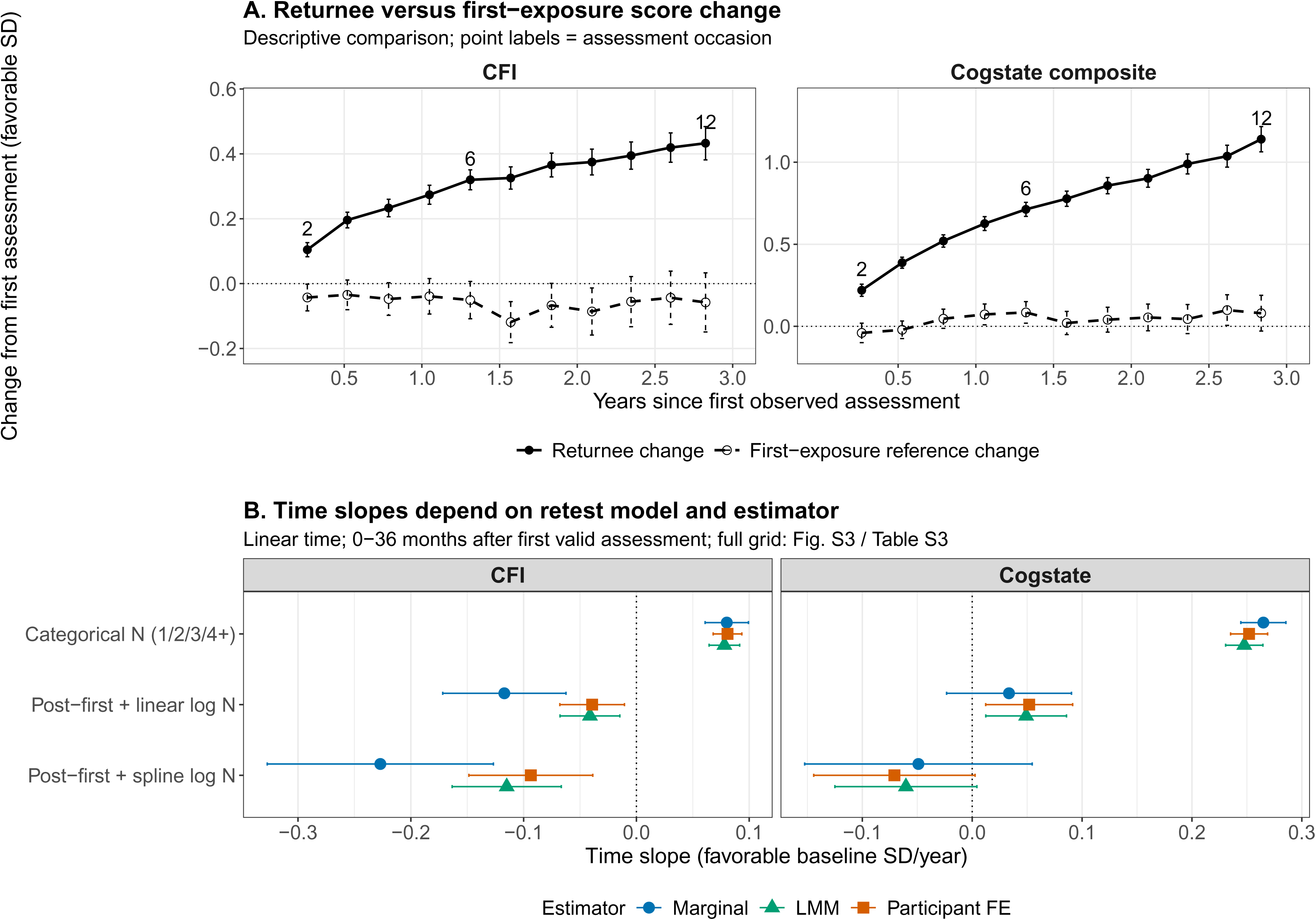
Observed score changes and the dependence of elapsed-time estimates on repeated-assessment specifications. **(A)** Mean change from the first observed assessment among repeatedly assessed participants (returnees) and the corresponding contrast derived from contemporaneously matched first-exposure observations. For each returnee assessment occasion, two distinct participants completing their first observed J-TRC assessment were matched separately to the returnee’s initial and later assessment dates, using calendar date, age, sex, and educational attainment. The first-exposure reference change represents the difference between these two cross-sectional observations, whereas the returnee change is calculated within an individual. Points represent estimates, and vertical intervals represent occasion-specific 95% confidence intervals. Labels 2, 6, and 12 identify selected assessment occasions, not sample sizes. Results are expressed in favorable baseline-standard-deviation units, with positive values indicating improvement. Full matched comparisons are reported in Supplementary Table S4A. **(B)** Estimated annual elapsed-time coefficients over the first 36 months after the first valid assessment of the respective instrument under three representative repeated-assessment specifications: categorical assessment number (1/2/3/4+), an indicator for assessments after the first combined with linear log assessment number, and an indicator combined with a spline of log assessment number. Estimates are shown for working-independence marginal regression (blue circles), maximum-likelihood linear mixed-effects models (green triangles), and participant fixed-effects models (orange squares). Horizontal intervals represent 95% confidence intervals. Positive coefficients indicate favorable change in baseline-standard-deviation units. The fixed-effects models require at least two observations per participant and therefore do not necessarily involve the same analytical population as the other estimators. The full specification grid is presented in Supplementary Figure S3 and Supplementary Table S3; sensitivity to the time origin is reported in Supplementary Table S5. The first-exposure reference is a matched between-participant contrast, and the elapsed-time coefficients are conditional on the specified retest models. **Abbreviations:** CFI, Cognitive Function Instrument; CI, confidence interval; FE, fixed effects; J-TRC, Japanese Trial-Ready Cohort; LMM, linear mixed-effects model; SD, standard deviation.

We summarized occasion-specific mean changes, reference contrasts, and their differences, with 95% confidence intervals (Supplementary Table S4A). An overall reference slope was estimated using participant-equal-weighted regression through the origin. Confidence intervals for this slope were obtained from 500 participant-level bootstrap replicates with repeated matching. Sensitivity analyses restricted first-exposure assessments to within 90 days of registration or narrowed the calendar matching window to ±90 days (Supplementary Figure S4; Supplementary Table S4B). The comparator pool could include participants registering after the primary March 2025 cutoff. The reference contrasts compared different participants at their first recorded J-TRC assessments, and previous cognitive testing outside J-TRC could not be assessed.

We examined the dependence of elapsed-time estimates on retest specification using assessments collected within 36 months of each participant’s first valid assessment of the respective instrument (Figure 4B). For each instrument, the latest analyzable assessment within each nonoverlapping 90-day interval anchored to registration was retained. Five representations of cumulative assessment exposure were evaluated, including categorical assessment numbers and linear or spline functions of log assessment number. Each specification included the interval since the preceding assessment and either linear elapsed time or a three-degree-of-freedom natural spline of elapsed time. Models adjusted for baseline age, sex, educational attainment, and registration year. Elapsed time was measured in years since the first valid assessment, whereas baseline age was defined at registration.

For each specification, we compared working-independence marginal Gaussian regression with participant-clustered robust inference, maximum-likelihood linear mixed-effects models, and participant fixed-effects models. Mixed-effects models included participant-specific random intercepts and time slopes where supported by the data, whereas fixed-effects models required at least two observations per participant. Linear-time coefficients were reported per year, and spline-time estimates were summarized using the model-implied 0–36-month contrast divided by three years. All coefficients were expressed in favorable baseline-standard-deviation units. Model fit was compared using AIC within comparable marginal and mixed-effects models, and predictive performance was evaluated using five repetitions of participant-clustered 10-fold cross-validation for marginal models. The full specification grid is provided in Supplementary Table S3. These analyses quantified the sensitivity of elapsed-time estimates to retest specifications and statistical estimators.

Sensitivity to selective observation was examined using inverse-probability-of-observation weighting (IPOW) across instrument-specific rolling risk intervals [Seaman2013(PMID: 21220355); Cole2008(PMID: 18682488)] (Supplementary Figure S5; Supplementary Table S4C). Interval-specific stabilized weights came from logistic observation models: numerator models included elapsed time and baseline covariates; denominator models additionally included previous scores, cumulative assessment history, recent activity, and lagged observation. The first assessment received weight one, and interval weights were not accumulated across previous intervals. We assessed 1st–99th percentile and alternative truncation, observation probabilities, weight distributions, and effective sample sizes. Complete-case outcome models did not impute unobserved scores. We also evaluated alternative time origins using the same assessment records (Supplementary Table S5).

Analyses were performed in R using data.table, survival, lme4, and related packages. Two-sided 95% confidence intervals were calculated using participant-clustered inference or bootstrap resampling as specified for each analysis.

## Results

### Registry growth and longitudinal data yield

Between October 2019 and the registration cutoff of March 31, 2025, the J-TRC Webstudy accumulated 14,912 registered accounts, of which 12,068 (80.9%) had provided explicit affirmative consent and 10,863 met the eligibility criteria (Figure 2A,B; Table 1). Among eligible participants, the mean age was 62.5 years (SD 8.4), 54.6% of those with recorded sex were women, and the mean educational attainment was 15.2 years (SD 2.0). Registration increased substantially during 2022, which accounted for 4,326 (39.8%) of eligible participants. A total of 10,645 eligible participants (98.0%) completed at least one valid assessment, all of whom had CFI data; 6,419 (59.1%) completed at least one valid Cogstate assessment. Overall, 5,792 participants (53.3%) contributed assessments on at least two distinct days.

Longitudinal data contribution decreased substantially with increasing time since registration (Table 1; Supplementary Table S2). At least one assessment at or beyond 12 months was available from 3,611 of 10,863 eligible participants (33.2%). Among participants with sufficient observation opportunity, 1,936 of 9,977 (19.4%) contributed data at or beyond 36 months and 651 of 4,602 (14.1%) at or beyond 60 months. The corresponding proportions for CFI were 33.1%, 19.3%, and 14.0%, compared with 26.8%, 16.9%, and 12.9% for Cogstate. Only 1,825 eligible participants (16.8%) had completed a valid assessment within the 365 days preceding the administrative observation cutoff of May 31, 2026. Thus, high initial assessment completion was accompanied by substantially lower rates of long-term data contribution.

Participation patterns were similar across 2019–2021 (n=5,133) and 2022–2023 (n=5,212) registration cohorts during a common 24-month window (Supplementary Tables S1 and S2). Initial CFI completion was 98.1% versus 97.9%, repeated CFI completion 51.8% versus 51.9%, initial Cogstate completion 60.4% versus 57.8%, and data contribution beyond 12 months 34.9% versus 32.3%.

Descriptive comparisons across participation stages showed differences in measured participant characteristics, particularly educational attainment and sex. Compared with participants without a valid Cogstate assessment, those completing a valid Cogstate assessment had higher educational attainment (standardized mean difference [SMD], 0.242) and a lower proportion of women (SMD, −0.219). Across the examined contribution-stage comparisons, absolute SMDs did not exceed 0.242.

### Repeated assessments and observed score distributions

Contributing participant numbers decreased across assessment occasions (Figure 2C): for CFI, 10,645 completed occasion 1, 5,610 occasion 2, 2,072 occasion 9, and 435 occasion 21; for Cogstate, 6,419, 3,994, and 384 completed occasions 1, 2, and 21. Occasions were not scheduled visits; corresponding elapsed times varied across participants (Supplementary Figure S1).

Observed score distributions also shifted across assessment occasions (Figure 2D,E). The median CFI total score, for which higher values indicate greater self-reported cognitive-functional difficulties, was 3.0 (interquartile range [IQR], 1.5–5.0) at the first assessment, 2.5 (IQR, 1.5–4.5) at the second, and 1.5 (IQR, 0.5–3.5) at the 21st. The median Cogstate composite score increased from 0.04 (IQR, −0.34 to 0.39) at the first assessment to 0.21 (IQR, −0.15 to 0.54) at the second and 0.83 (IQR, 0.52–1.20) at the 21st. These distributions summarize the participants observed at each assessment occasion, whose composition changed over time.

### Dynamic participation, inactivity, and selective observation

Participation status changed markedly with time since registration (Figure 3A; Supplementary Table S2). At approximately 1.23 years after registration, 5,375 of 10,833 participants with sufficient observation opportunity (49.6%) had completed an assessment within the preceding 365 days, whereas 5,208 (48.1%) had previously been assessed but were inactive for at least 365 days. At approximately 3.20 years, the proportion classified as inactive had increased to 75.7% (7,279/9,617), and by 6.16 years it reached 83.5% (2,348/2,813). The Kaplan–Meier median time to the first assessment gap of at least 365 days was 1.00 year for CFI and 1.37 years for Cogstate; the probabilities of remaining without such a gap at three years were 20.3% and 29.6%, respectively (Supplementary Table S2).

Prolonged inactivity was not invariably followed by permanent cessation of assessment activity (Figure 3B). Following the first period of at least 365 consecutive days without a valid assessment, the Kaplan–Meier cumulative probabilities of reactivation were 3.6% (95% CI, 3.3–4.0%) at approximately six months, 5.2% (95% CI, 4.8–5.7%) at 12 months, and 7.2% (95% CI, 6.7–7.8%) at 24 months. The corresponding numbers at risk were 8,456, 7,983, and 6,896. Estimates varied with the operational definition of prolonged inactivity: when inactivity was defined using a 180-day threshold, cumulative reactivation at 24 months was 13.1%, compared with 3.5% using a 730-day threshold (Supplementary Figure S2; Supplementary Table S2). Among 666 reactivation events with at least 365 days of subsequent observation opportunity, 203 (30.5%) were followed by assessment activity on at least two additional days within the following year.

Subsequent observation was associated with both prior assessment scores and recent participation history (Figure 3C,D; Supplementary Table S2). In adjusted models of 168,263 CFI risk intervals from 9,611 participants, each 1-SD worse prior CFI score was associated with lower odds of a subsequent observed assessment (odds ratio [OR], 0.952; 95% CI, 0.929–0.976; cluster-robust *P* < 0.001). The corresponding association for Cogstate was stronger (OR, 0.845; 95% CI, 0.819–0.872; *P* < 0.001), based on 102,479 intervals from 5,894 participants. Model-standardized probabilities of subsequent observation also differed according to time since the most recent Webstudy assessment activity, whether CFI or Cogstate. For CFI, the predicted probability decreased from 22.8% for a recency category of less than 90 days to 13.1% for 730 days or longer; the corresponding values for Cogstate were 34.3% and 2.6%. These findings indicate that subsequent assessment completion was associated with previously observed scores and assessment histories.

### Interpretation of longitudinal score changes

Among participants who returned for repeated assessments, observed score changes were consistently more favorable than the corresponding contrasts derived from contemporaneous first-observed J-TRC assessments (Figure 4A; Supplementary Table S4A). By the 12th assessment occasion, the estimated change from the first assessment among matched returnees was +0.433 favorable baseline SD (95% CI, 0.381–0.484) for CFI and +1.140 SD (95% CI, 1.063–1.217) for Cogstate. The corresponding first-exposure reference contrasts were −0.058 SD (95% CI, −0.149 to 0.033) and +0.080 SD (95% CI, −0.029 to 0.189), yielding differences of +0.491 SD (95% CI, 0.385–0.596) and +1.060 SD (95% CI, 0.925–1.196), respectively. At this occasion, 931 CFI returnees (83.7% of eligible returnees) and 628 Cogstate returnees (67.8%) were successfully matched. Across the matched observations, the participant-equal first-exposure reference slopes were −0.033 favorable SD/year for CFI (95% rematching-bootstrap CI, −0.057 to −0.007) and +0.025 SD/year for Cogstate (95% CI, −0.005 to 0.054). Restricting the reference to participants whose first J-TRC assessment occurred within 90 days of registration yielded similar estimates of −0.038 SD/year (95% CI, −0.058 to −0.009) and +0.021 SD/year (95% CI, −0.004 to 0.058), respectively (Supplementary Figure S4; Supplementary Table S4B). These comparisons quantify differences between within-person returnee changes and matched first-exposure contrasts from different participants.

The estimated elapsed-time associations varied substantially depending on how repeated-assessment exposure was represented (Figure 4B; Supplementary Figure S3; Supplementary Table S3). In working-independence marginal models over the first 36 months after the first valid assessment, categorical adjustment for assessment number (1/2/3/4+) yielded annual coefficients of +0.080 favorable baseline SD/year for CFI (95% CI, 0.061–0.099) and +0.265 SD/year for Cogstate (95% CI, 0.245–0.285). With a post-first-assessment indicator and a spline of log assessment number, the corresponding coefficients were −0.227 SD/year (95% CI, −0.327 to −0.127) and −0.049 SD/year (95% CI, −0.153 to 0.055).

Similar dependence on retest specification was observed with mixed-effects and participant fixed-effects models. For CFI, the categorical and spline-log specifications yielded coefficients of +0.078 versus −0.115 SD/year in mixed-effects models and +0.081 versus −0.094 SD/year in fixed-effects models. The corresponding Cogstate estimates were +0.248 versus −0.060 SD/year and +0.252 versus −0.071 SD/year, respectively. Among linear-time models, AIC favored more flexible retest representations. Across marginal models with linear or spline elapsed-time functions, cross-validated prediction errors differed little; the maximum excess root mean squared error was 0.125% for CFI and 0.377% for Cogstate. Thus, substantial variation in elapsed-time estimates was accompanied by only small differences in cross-validated prediction error among marginal models. Sensitivity analyses using time since registration or additionally adjusting for the delay from registration to the first valid assessment are reported in Supplementary Table S5.

Inverse-probability-of-observation weighting sensitivity analyses remained dependent on retest specification and weight truncation (Supplementary Figure S5; Supplementary Table S4C). Estimated observation probabilities were below 0.01 for 49.9% of CFI intervals and 43.4% of Cogstate intervals, while effective sample sizes under the primary weighting scheme were only 13.3% and 26.8% of observed weighted assessments, respectively.

## Discussion

Using over six years of J-TRC Webstudy data, we found high initial assessment completion but limited multiyear contribution; frequent inactivity, occasional reactivation, and score-associated subsequent observation; and more favorable returnee changes than first-exposure contrasts. In marginal models, estimated elapsed-time coefficients varied markedly across retest specifications despite similar cross-validated prediction errors. Together, these findings show the importance of sustained participation, selective observation, and retest-model specification when interpreting longitudinal cognitive scores from web-based registries.

The Brain Health Registry and Alzheimer Prevention Trials Webstudy have demonstrated large-scale remote recruitment [Weiner2018(PMID: 29754989); Walter2020(PMID: 32920623)], while other studies have examined unsupervised cognitive assessment [Banh2022(PMID: 35542999)]. Early J-TRC recruitment strategies and their short-term effects have been reported previously [Sato2021(PMID: 34292159)], as have Webstudy usability and questionnaire participation [Sato2024(PMID: 38706282); Nakashima2025(PMID: 40115874)]. Because assessment schedules and follow-up definitions differ, retention rates cannot be directly compared across registries. Our study extends this literature by characterizing opportunity-adjusted multiyear data contribution, inactivity and reactivation, score-associated subsequent observation, and the model dependence of repeated cognitive score interpretation.

Longitudinal data contribution was concentrated early: almost all eligible participants completed CFI initially, approximately half contributed on multiple days, and far fewer contributed after three or five years. These proportions are not conventional fixed-visit retention rates because assessments were voluntary and observation opportunity differed. Inactivity did not necessarily mean withdrawal: reactivation within two years after a 365-day gap occurred with 7.2% cumulative probability, sometimes followed by further assessments. Similar 24-month patterns across registration cohorts suggest that reduced longer-term contribution was not confined to early recruitment. Registries should distinguish activity, inactivity, and return, and report recent activity separately from longer-term contribution. Neither directly establishes readiness for an onsite study or trial.

The association between prior cognitive scores and subsequent observation has additional implications for the interpretation of longitudinal data. Participants with less favorable prior scores were less likely to complete subsequent assessments, particularly for Cogstate, while recent assessment activity was strongly associated with further observation. Consequently, participants contributing data at later assessment occasions represented a progressively selected subset of the original eligible cohort. The occasion-specific score distributions observed in this study therefore cannot be interpreted as longitudinal trajectories of the full registry population. Similar concerns have been raised in statistical research on informative observation processes, in which the probability of obtaining an outcome measurement is related to observed or unobserved participant characteristics [Gasparini2020(PMID: 31894164); Pullenayegum2016(PMID: 24855119)]. These findings demonstrate that subsequent assessment was associated with prior cognitive scores and participation histories, highlighting the importance of the observation process when interpreting longitudinal cognitive data.

The favorable score changes among returnees were substantially greater than contemporaneous first-exposure contrasts. By assessment 12, favorable changes reached 0.43 baseline SD for CFI and 1.14 SD for Cogstate, whereas the first-exposure contrasts were close to zero. Several mechanisms may contribute to these differences, including repeated exposure, familiarity with testing, and selective continuation. Practice-related gains have been documented in longitudinal cognitive research [Salthouse2010(PMID: 20804244); Hoffman2011(PMID: 21639642); Chen2023(PMID: 37199336)]. Recent randomized evidence also showed that the number and frequency of previous Cogstate assessments influence subsequent performance [Murchland2026(PMID: 42224199)]. For the self-reported CFI, changes in symptom reporting [Shrout2018(PMID: 29255039)] and the handling of non-scorable items may also contribute. Together, these findings highlight the importance of assessment exposure and changing participant composition when interpreting longitudinal online cognitive scores.

Elapsed-time coefficients varied materially across retest specifications. This dependence persisted when CFI analyses were restricted to fully scorable responses or used prorated totals (Supplementary Table S1). With linear time, AIC favored more flexible retest models in marginal and mixed-effects analyses (mixed-effects AIC differences: 76.1 for CFI; 99.0 for Cogstate), but participant-clustered cross-validated RMSE was nearly unchanged. Categorical occasion adjustment gave favorable time coefficients, whereas flexible specifications yielded less favorable or negative estimates across marginal, mixed-effects, and fixed-effects models. Estimators also used different sources of variation and, for fixed effects, a restricted population (Supplementary Table S3). Time and cumulative assessment count were strongly correlated, limiting their empirical separation [Ferrer2004(PMID: 15222818); Hoffman2011(PMID: 21639642); Vivot2016(PMID: 26825924)]. The marked variation in elapsed-time estimates despite similar prediction errors highlights the importance of retest-model specification for interpreting longitudinal cognitive change.

The contemporaneous first-exposure comparisons provided an empirical reference for observed returnee changes. Constraining elapsed-time coefficients to the reference slopes had little effect on cross-validated prediction error (Supplementary Table S3). IPOW and alternative time-origin analyses further showed that elapsed-time estimates remained sensitive to retest specification, while low observation probabilities and reduced effective sample sizes limited weighted inference (Supplementary Figure S5; Supplementary Tables S4C and S5). Together, these findings identified retest-model specification as an important source of variation in estimated longitudinal trends.

These results have practical implications for trial-ready registries. Reports should separate enrollment, eligibility, recent activity, and multiyear data contribution using opportunity-adjusted denominators. Assessment dates, cumulative exposure, intervals, inactivity, and reactivation should be preserved for analysis. Future studies should test whether reminder timing and frequency improve long-term assessment participation. Longitudinal modeling should report sensitivity to plausible retest specifications, time origins, and estimators, rather than a single adjusted trajectory. Repeated online scores also require validation against independently measured cognitive or biomarker outcomes before use in natural-history estimation or risk enrichment.

In J-TRC, A4-derived models incorporating CFI and Cogstate scores have been used to prioritize onsite invitations [Sato2021(PMID: 33778148); Nakashima2026(PMID: 41536849)]. One A4-derived model included a score difference between screening visits 1 and 3 [Sato2021(PMID: 33778148)], whereas Webstudy participants could complete tests over longer, irregular intervals. Whether this heterogeneity affects amyloid-risk predictions or invitation priority was not assessed. ADNI-3 findings demonstrate longitudinal utility of remote Cogstate under structured follow-up [Maruff2026(PMID: 42138182)]. More regular schedules, prospectively defined first-exposure comparators, documentation of test versions, and linkage to clinical or biomarker outcomes warrant evaluation. The AlzMatch pilot additionally showed feasibility of remote registry-linked blood collection [Walter2024(PMID: 39350391)]. Online scores are informative, but should not be treated as equivalent to scheduled clinical follow-up.

This study has several strengths, including its large nationwide registry population, more than six years of observation, explicit accounting of registration and eligibility denominators, and repeated assessment data from both CFI and Cogstate. The availability of assessment dates and histories allowed us to evaluate long-term data contribution, inactivity, reactivation, score-associated subsequent observation, and sensitivity to retest-model specification within the same cohort.

Several limitations should be considered. Participants were self-selected internet users with relatively high educational attainment, and eligibility was partly self-reported, limiting generalizability [Hall2015(PMID: 25156311); Ashford2020(PMID: 32478165)]. Cogstate assessment was optional, and experiences with unsupervised cognitive testing may differ across participants [Ashford2023(PMID: 37357303)]. Assessment timing was not randomized, the reasons for stopping or resuming assessments were unknown, and cognitive scores during unobserved periods were unavailable. Thus, we could not determine the direction or magnitude of bias related to selective observation. First-exposure reference contrasts compared different participants matched on a limited set of measured characteristics, leaving possible differences in unmeasured factors, calendar time, and the composition of new registrants. These contrasts do not isolate causal effects of repeated testing or individual cognitive aging trajectories. Fewer participants contributed data at later assessment occasions. Some models required complete baseline covariates or repeated observations, resulting in analytic samples that differed from the eligible cohort. Those excluded because of incomplete covariate information were more often women (Supplementary Table S1). IPOW analyses were additionally limited by very low estimated observation probabilities and small effective sample sizes. Finally, we did not directly validate longitudinal scores against independently adjudicated clinical progression or AD biomarkers. These limitations should be considered when interpreting the observed cognitive score patterns.

## Conclusions

The J-TRC Webstudy achieved high initial assessment completion in a large nationwide cohort, while sustained data contribution became concentrated among a smaller group. Subsequent assessment was associated with prior scores and participation history, and returnee changes were substantially more favorable than contemporaneous first-exposure contrasts. Estimated elapsed-time coefficients differed markedly across retest specifications. These findings demonstrate the complementary importance of opportunity-adjusted data yield, assessment history, and retest-model sensitivity for evaluating web-based registries and interpreting longitudinal cognitive scores in AD research.

## Data Availability

The J-TRC Webstudy data supporting the findings of this study are not publicly available due to ethical restrictions.

## Acknowledgements

Some authors’ affiliation (KS, YN, TI), “*Dementia Inclusion and Therapeutics*,” is an endowed department at the University of Tokyo Hospital funded by Effissimo Capital Management Pte Ltd. AI-assisted tools were used for language editing; authors take full responsibility for the content.

## Consent Statement

All participants provided online informed consent through the Webstudy registration process.

