## Supplementary Materials for "Longitudinal Participation and Cognitive Score Interpretation in the J-TRC Webstudy"

**Supplementary Methods**

**S1. Reconstruction of the registry population and analytical denominators**

The registry population was reconstructed using registration records, electronic consent status, self-reported eligibility information, and dates of valid cognitive assessments. The primary registration cutoff was March 31, 2025, and the administrative observation cutoff was May 31, 2026. Registered accounts were distinguished from participants providing explicit affirmative consent. Advancement through the registration workflow was not considered sufficient evidence of consent in the absence of an affirmative consent response.

Eligibility was determined independently of whether participants had completed the final registration workflow or completed valid cognitive assessments. The eligible consented cohort comprised participants aged 50–85 years at registration who had an affirmative consent response, ascertainable age and self-reported Alzheimer's disease (AD) status, and no self-reported previous AD diagnosis. Participants with unavailable age or AD status were classified as having insufficient information to establish eligibility rather than being considered ineligible. Completion of the final information-registration workflow was evaluated separately as a secondary administrative measure.

For analyses of assessment activity, a valid CFI or Cogstate assessment defined an observed assessment event. When both instruments were completed on the same calendar day, the combined activity measure counted one activity day. Repeated participation required activity on at least two distinct calendar days. Contributions at or beyond 12, 36, and 60 months were assessed relative to individual registration dates. The denominator for each threshold was restricted to participants whose registration date preceded the administrative cutoff by at least the corresponding interval. Participants who had not yet accumulated sufficient observation opportunity were excluded from that threshold-specific denominator, rather than classified as noncontributors.

**S2. Assessment processing, score construction, and quality control**

**CFI processing.** The 15-item Cognitive Function Instrument (CFI) was scored using item responses of "No" (0), "Maybe" (0.5), and "Yes" (1). A response of "Does not apply" was accepted as an answered item for determining questionnaire completeness but was excluded from the numerical item sum. A valid primary assessment required a valid assessment date, an available recorded total, and valid response categories for all 15 items. The primary score was the sum of scorable item values without prorating for "Does not apply" responses. Higher scores indicated more self-reported cognitive-functional difficulties.

Recorded CFI totals were independently reconstructed from item responses to check scoring consistency. Exact duplicate records were removed, and where more than one valid CFI record was available for an individual on the same calendar day, the latest record was retained. Scoring sensitivity analyses were conducted using (i) assessments for which all 15 items were numerically scorable and (ii) totals prorated to 15 items based on the number of scorable responses. These alternatives were evaluated separately from the primary scoring convention (Supplementary Table S1).

**Cogstate processing.** Four exported domain scores were used: Detection, Identification, One Card Learning, and One Back. Exact duplicate sessions were removed. Each domain was standardized using the mean and standard deviation of that domain at participants' first observed Cogstate assessments. The available standardized domain values were averaged to construct the composite score. Although the processing algorithm permitted composite calculation when at least two domain values were available, all 40,058 sessions retained in the primary analytic export contained the four prespecified domain values.

The analytic pipeline treated the exported domain values as higher-is-better scores.

**Standardized change scores.** For comparisons of longitudinal change, each outcome was scaled by its standard deviation at the first observed assessment. The sign of CFI change was reversed so that positive values indicated fewer reported difficulties; positive values for the Cogstate composite indicated higher performance under the analytic scoring convention. Consequently, favorable standardized coefficients were used for comparisons across outcomes, whereas descriptive CFI totals and selected sensitivity analyses retained their original scoring units.

**S3. Assessment occasions, registration cohorts, and participant composition**

Assessment occasion was defined separately for each instrument as the ordinal position of an individual's valid assessments. It was not assigned according to a common visit schedule. The number of contributing participants and the observed median and interquartile range were calculated for each occasion. To examine the support for distinguishing elapsed time from cumulative exposure, we also summarized elapsed time since registration by assessment number and calculated Pearson and Spearman correlations, together with variance inflation factors from auxiliary time-exposure models (Supplementary Figure S1).

The consistency of participation patterns across enrollment periods was assessed by comparing registrations from 2019–2021 with registrations from 2022–2023. Both cohorts were evaluated over a common 24-month observation window to reduce differences caused by unequal potential follow-up. Outcomes included initial valid assessment completion, repeated assessment, and contribution beyond 12 months.

Measured characteristics were compared between participants who did and did not reach selected contribution stages. Standardized mean differences (SMDs) were calculated for age, sex, educational attainment, and registration-year indicators. These comparisons were descriptive and were not intended to estimate causal or independent determinants of participation (Supplementary Tables S1 and S2).

**S4. Time-dependent participation states, prolonged inactivity, and reactivation**

Participant activity states were evaluated on a registration-anchored grid of nonoverlapping 90-day intervals. At each interval endpoint, participants with sufficient administrative observation opportunity were classified into three mutually exclusive states: no previous valid assessment, recently active, or prolonged inactivity. Recently active indicated assessment activity during the preceding 365 days; prolonged inactivity indicated previous assessment activity followed by at least 365 consecutive days without a valid assessment. Incomplete terminal intervals were excluded.

This registration-anchored state analysis was distinguished from the rolling instrument-specific availability intervals used for the subsequent-observation regression models.

For time-to-event analyses, the first qualifying prolonged-inactivity gap was identified separately for CFI and Cogstate, beginning after the instrument's first observed assessment. Time to the first interval of at least 365 consecutive days without assessment was analyzed using Kaplan–Meier estimation. Participants who did not satisfy the inactivity criterion were censored at the administrative observation cutoff.

Reactivation was defined as a subsequent valid assessment after the first qualifying prolonged-inactivity period. Follow-up for the reactivation analysis began when the inactivity threshold was attained and continued until the next valid assessment or administrative censoring. Cumulative reactivation probabilities and corresponding 95% confidence intervals were obtained using Kaplan–Meier methods. Numbers at risk were determined at prespecified follow-up times. Among reactivation events with at least 365 days of subsequent observation opportunity, additional engagement was assessed by counting distinct activity days during the following year.

Sensitivity analyses used inactivity thresholds of 180 and 730 days in place of 365 days. Registration-anchored activity-state prevalence was also recalculated using panel widths of 60 and 120 days instead of 90 days (Supplementary Figure S2). Results under different inactivity thresholds were considered distinct operational estimands because the definitions changed both the qualifying events and the populations at risk.

**S5. Rolling assessment-availability intervals and models of subsequent observation**

For each outcome, we constructed rolling assessment-availability intervals to evaluate whether a subsequent valid assessment was observed after reassessment became available. The observation process was modeled using complete risk intervals, taking account of the 90-day interval after the previous assessment during which reassessment of the same instrument was not available. Terminal intervals without complete opportunity for observing the outcome were excluded.

Interval-specific observation was modeled using logistic regression. Candidate predictors included time since registration and its squared term, age, sex, educational attainment, registration year, the most recently observed instrument score, cumulative assessment count, recent assessment activity, and previous observation indicators. Prior-score history was expressed in standard-deviation units, with its direction aligned so that higher values represented less favorable performance. Missing prior-score information was represented by a missingness indicator together with median imputation where required for prediction.

Associations between prior score and subsequent assessment observation were reported as odds ratios for a 1-SD worse prior score. Confidence intervals used participant-clustered sandwich estimators to account for repeated risk intervals within individuals. Model-standardized probabilities were calculated by averaging predicted probabilities across the empirical distribution of covariates while varying the predictor of interest. Time since previous activity was additionally summarized using categories of less than 90, 90–179, 180–364, 365–729, and at least 730 days. These models described observed associations rather than causal mechanisms of disengagement.

**S6. Contemporaneous first-exposure comparison and bootstrap inference**

To provide an empirical comparison for changes among repeatedly assessed participants, we constructed an internal contemporaneous first-exposure reference for CFI and Cogstate separately. Eligible returnee assessments were restricted to occasions 2–12 occurring within 36 months of the returnee's first observed assessment.

For each returnee and assessment occasion, the difference between the repeated and initial scores constituted the within-person returnee change. Two first-exposure comparator observations were then selected separately: one corresponding to the returnee's initial assessment and another corresponding to the returnee's later assessment. The resulting reference contrast was the difference between the later and earlier first-exposure comparator scores. The two comparator observations came from different participants and therefore did not constitute an individual longitudinal trajectory (Supplementary Figure S4A).

Matching was performed using a greedy nearest-neighbor algorithm, without replacement within each matching analysis. Eligibility required the same sex, calendar dates within ±180 days, attained ages within ±2 years, and educational attainment within ±2 years. The same first-exposure comparator participant was not reused in both the baseline and later comparator roles for a matched returnee contrast. Matching was conducted separately for each assessment occasion and outcome.

For each occasion, we summarized eligible returnees, matched returnees, median elapsed time, the within-person returnee change, the first-exposure reference contrast, and their difference. Occasion-specific confidence intervals were calculated separately from the rematching-bootstrap procedure used for the overall reference slope.

The reference slope was estimated by regression of matched first-exposure contrasts on returnee elapsed time, constrained to pass through the origin, with equal total weight assigned to each returnee participant. Uncertainty was assessed using participant-level bootstrap resampling, repeating the matching procedure within each replicate. Percentile confidence intervals were obtained from the bootstrap distribution; the number of valid replicates was recorded explicitly.

Sensitivity analyses included restricting comparator first exposures to those occurring within 90 days after registration and narrowing the calendar matching window to ±90 days. The restriction to first assessments within 90 days of registration used 500 participant-level bootstrap replicates with rematching, whereas the narrower-calendar-window analysis used participant-clustered robust confidence intervals. These intervals were not treated as interchangeable. We also examined measured differences between earlier and later first-exposure entrants using SMDs.

The comparator pool was not restricted to the primary March 2025 registration cutoff, allowing first-exposure assessments from later registrants before the administrative observation cutoff. First observed exposure in J-TRC did not imply lifelong absence of prior cognitive testing. Neither the resulting reference slope nor its comparison with returnees was interpreted as a biological aging trajectory or a causal estimate of repeated-testing effects.

**S7. Elapsed-time and retest-model specification grid**

Longitudinal model comparisons were restricted to assessments occurring within 36 months after each participant's first valid assessment of the respective instrument. For each outcome, the latest valid assessment within each registration-anchored 90-day interval was retained. Elapsed time in the longitudinal models was defined as years since the first valid assessment, with time zero corresponding to assessment number 1. Baseline age was defined at registration. A common complete-case analysis sample was used across the alternative retest representations within each outcome, except that participant fixed-effects models required at least two observations per participant.

Five representations of repeated-assessment exposure were evaluated:

1. Assessment number categorized as 1, 2, 3, or 4 and above.
2. An indicator for assessments after the first combined with a linear term in log assessment number.
3. Expanded assessment-number categories, distinguishing occasions 1–6, grouping occasions 7–9, and grouping occasion 10 and above.
4. Individual indicators through assessment number 10, with subsequent occasions grouped as 11 and above.
5. An indicator for assessments after the first combined with a natural cubic spline of log assessment number with three degrees of freedom.

All specifications included log(1 + interval in months) to describe the interval since the preceding assessment; the first assessment was assigned an interval value of zero for this variable.

Each retest representation was fitted using either linear elapsed time or a natural cubic spline of elapsed time with three degrees of freedom. Baseline age, sex, educational attainment, and registration year were included as covariates.

Three estimators were compared: working-independence marginal Gaussian regression with participant-clustered robust inference; maximum-likelihood linear mixed-effects modeling, allowing participant-specific intercepts and time slopes when supported; and participant fixed-effects estimation. The resulting grid comprised 30 models per outcome, or 60 models overall.

For linear-time models, the time-associated estimate was the coefficient per year. For spline-time models, an annualized summary was obtained from the model-predicted contrast between months 0 and 36, divided by three years. In both cases, months 0 and 36 were defined relative to the first valid assessment, rather than registration. All reported coefficients were converted to favorable baseline-SD units.

Model fit was compared using AIC within outcome, time-function, and eligible estimator class. AIC differences were not used to rank fundamentally different estimators, and no corresponding fixed-effects AIC comparison was reported. Predictive performance for marginal models was evaluated using five repetitions of participant-clustered 10-fold cross-validation, with folds formed at the participant level. Root mean squared error (RMSE) was calculated for change-from-first predictions in native outcome units. As an exploratory sensitivity analysis, we refitted linear-time marginal models with the elapsed-time coefficient constrained to the separately estimated first-exposure reference slope while re-estimating the remaining model terms. The reference slope was applied as an offset using elapsed time since the first valid assessment, such that the offset was zero at the first assessment. Participant-clustered cross-validated RMSE was compared with that from the unconstrained models. This analysis evaluated prediction sensitivity to the numerical constraint and did not assume that the reference contrast and longitudinal coefficient estimated the same underlying quantity.

We evaluated sensitivity to the time origin using the same first-assessment-anchored 36-month analysis sample. Linear-time models were fitted using (1) elapsed time since the first valid assessment (primary analysis), (2) elapsed time since registration, and (3) elapsed time since the first valid assessment with additional adjustment for the interval between registration and the first valid assessment. The same three retest specifications and three estimator classes shown in Figure 4B were compared (Supplementary Table S5). These analyses evaluated sensitivity to the definition of elapsed time without changing the observation window or analytical sample.

**S8. Observation weighting and selective-follow-up sensitivity analyses**

Inverse-probability-of-observation weighting (IPOW) was used to investigate the sensitivity of time-associated estimates to the observed assessment process. Stabilized interval-specific weights were constructed from logistic models of whether an assessment was observed during an eligible rolling risk interval.

The numerator model included time and baseline covariates, whereas the denominator model additionally incorporated available time-varying information, including prior scores, cumulative assessment history, recent activity, and lagged observation indicators. The first observed assessment was assigned a weight of one. Interval-specific stabilized weights were not multiplied cumulatively across previous observed and missed assessment opportunities.

Primary weights were truncated at the 1st and 99th percentiles of the observed weight distribution. Sensitivity analyses used truncation at the 2.5th–97.5th and 5th–95th percentiles, alternative 60- and 120-day rolling risk intervals, and more flexible functional forms for the observation model. We summarized the distribution of estimated observation probabilities, the proportion below 0.01, the percentage of truncated weights, and the effective sample size, calculated as the squared sum of weights divided by the sum of squared weights.

Additional analyses examined time coefficients under alternative retest specifications and among participants with at least 36 months of potential follow-up, stratified by their observed participation histories. These analyses assessed sensitivity to observation and retention patterns; they were not assumed to identify the distribution of unobserved cognitive outcomes.

IPOW outcome coefficients were retained in native units—CFI points per year or Cogstate composite z-score units per year—in Supplementary Table S4C. Standardized favorable-direction coefficients were used for the graphical comparison in Supplementary Figure S5. Confidence intervals conditioned on the estimated weighting procedure and should not be interpreted as accounting for all uncertainty associated with the observation models.

**S9. Missing data and reproducibility considerations**

Missing educational attainment or other baseline covariates led to exclusion from complete-case regression models requiring those variables. Descriptive analyses and eligibility definitions did not require complete covariate information unless explicitly stated. Additional analyses retaining participants with missing education through an explicit missingness category were used to assess sensitivity of selected longitudinal coefficients. Unobserved cognitive outcomes were not imputed in the main analysis.

The analytical pipeline was implemented in R using data.table, survival, lme4, and associated packages. The final reference analyses used 500 valid bootstrap estimates for each outcome under both the primary matching specification and the restriction to first assessments within 90 days of registration.

**Supplementary Results**

**S1. Cohort reconstruction, assessment availability, and scoring quality control**

Among 14,912 registered accounts, 12,068 had an explicit affirmative consent response. Age and self-reported AD status were ascertainable for 11,901 consented accounts (98.6%), while 167 (1.4%) had insufficient information to establish eligibility. The primary eligible consented cohort comprised 10,863 participants, of whom 10,347 (95.2%) had also completed the final registration workflow. The latter administrative milestone was not required for inclusion in the primary cohort (Supplementary Table S1).

The analytic assessment datasets contained 51,384 valid CFI participant-days and 40,058 valid Cogstate sessions. Before the CFI participant-day deduplication step, recorded totals agreed exactly with independently reconstructed item sums in all 51,395 valid sessions, including 18,191 sessions with at least one "Does not apply" response. After deduplication, 18,188 of 51,384 valid CFI assessment days (35.4%) contained at least one such response. Their frequency increased from 28.7% at the first occasion to 38.3% at the fourth or later occasions. All 40,058 analyzable Cogstate sessions contained the four prespecified exported domain values, and no participant-days contained multiple distinct complete Cogstate sessions after exact-duplicate removal (Supplementary Table S1).

Alternative CFI scoring conventions did not eliminate the sensitivity of elapsed-time coefficients to retest representation. When analysis was restricted to assessments with all 15 items numerically scorable, the estimated native-score annual coefficient was −0.084 CFI points/year under categorical assessment-number adjustment and +0.391 points/year under the post-first plus spline-log specification. The corresponding coefficients using prorated CFI totals were −0.061 and +0.406 points/year, respectively. Thus, the difference between coarse and flexible retest adjustment was retained under both scoring alternatives (Supplementary Table S1).

**S2. Participant composition, covariate missingness, and registration-cohort consistency**

Measured characteristics differed across some participation stages, although absolute SMDs for the examined variables generally remained below 0.25 (Supplementary Table S1). Participants completing a valid Cogstate assessment had higher educational attainment than those without a valid Cogstate assessment (SMD, 0.242) and a lower proportion of women (SMD, −0.219). Educational attainment was also higher among those completing a valid CFI assessment (SMD, 0.205). For contributions extending beyond 36 months, differences in registration-year composition were evident, reflecting differences in both recruitment timing and observation opportunity.

The complete-case longitudinal model populations comprised 9,611 CFI participants and 5,898 Cogstate participants. Compared with those retained in the respective models, participants excluded because of incomplete baseline covariate information were more often women: 80.3% versus 51.8% for CFI and 78.9% versus 47.6% for Cogstate. Excluded participants were also somewhat younger. These differences indicate that complete-case longitudinal model populations were not identical in composition to the full assessment-completion populations (Supplementary Table S1).

Participation measures were broadly similar across the earlier and later registration cohorts within the common 24-month window (Supplementary Table S2). The 2019–2021 cohort included 5,133 participants, compared with 5,212 in the 2022–2023 cohort. Completion of at least one valid CFI assessment was 98.1% versus 97.9%, and repeated CFI assessment was 51.8% versus 51.9%, respectively. Completion of at least one valid Cogstate assessment was 60.4% versus 57.8%, and contribution beyond 12 months was 34.9% versus 32.3%. Thus, the primary pattern of high initial assessment completion followed by substantially lower longitudinal contribution was observed in both enrollment periods.

**S3. Relationship between elapsed time and cumulative assessment number**

The timing of repeated assessments was heterogeneous despite the minimum 90-day reassessment interval (Supplementary Figure S1). The cumulative number of completed assessments was strongly correlated with elapsed time in the same-sample longitudinal datasets. Pearson correlations were 0.961 for CFI and 0.953 for Cogstate, with corresponding Spearman correlations of 0.971 and 0.959. Auxiliary models assessing the dependence of elapsed time on cumulative assessment exposure and assessment interval yielded variance inflation factors of 15.2 for CFI and 12.2 for Cogstate. These findings indicate substantial overlap in the temporal information represented by elapsed time and cumulative assessment number.

**S4. Prolonged inactivity and sensitivity to the operational definition**

In instrument-specific Kaplan–Meier analyses, the median time to the first assessment gap of at least 365 days was 1.00 year for CFI and 1.37 years for Cogstate (Supplementary Figure S2A,B; Supplementary Table S2). The estimated probabilities of remaining without a qualifying gap were 49.9% (95% CI, 48.9–50.8%) for CFI and 59.9% (58.7–61.1%) for Cogstate at one year. At three years, these probabilities were 20.3% and 29.6%, respectively, declining further to 13.7% and 20.8% at five years.

The estimated prevalence of inactivity and subsequent reactivation depended on the duration used to define a qualifying gap (Supplementary Figure S2C–E). At 24 months after reaching the inactivity threshold, cumulative reactivation probabilities were 13.1% (95% CI, 12.5–13.8%) for a 180-day definition, 7.2% (6.7–7.8%) for the primary 365-day definition, and 3.5% (3.1–4.0%) for a 730-day definition. Corresponding 12-month probabilities were 10.8%, 5.2%, and 2.2%. The absolute probabilities differed because each threshold defined a different qualifying inactivity episode and risk population.

Recalculation of registration-anchored inactivity prevalence using panel widths of 60, 90, and 120 days yielded broadly similar temporal patterns (Supplementary Figure S2F). Across panel definitions, inactivity became increasingly common with time since registration, without evidence that the primary qualitative pattern was driven by the choice of a 90-day grid.

**S5. Matching support and robustness of the first-exposure reference**

The proportion of eligible returnees successfully matched at both time points varied across assessment occasions and outcomes (Supplementary Figure S4B; Supplementary Table S4A). For CFI, matching proportions ranged from 76.7% to 87.4% across occasions 2–12. The corresponding proportions for Cogstate ranged from 55.4% to 79.4%. At the 12th occasion, 931 of 1,112 eligible CFI returnees (83.7%) and 628 of 926 eligible Cogstate returnees (67.8%) were successfully matched. The median elapsed times at that occasion were 2.83 and 2.84 years, respectively.

Across assessment occasions, the primary first-exposure analysis included 24,456 matched CFI returnee-occasion observations from 4,835 distinct returnees and 16,866 Cogstate observations from 3,250 distinct returnees (Supplementary Table S4B). The first-exposure comparator pools also included first assessments from individuals registering after the March 2025 cutoff: 708 for CFI and 19 for Cogstate. The matched comparisons therefore used a broader calendar-time comparator population than the primary registry-level analysis.

The participant-equal first-exposure reference slopes were −0.033 favorable baseline SD/year (95% bootstrap CI, −0.057 to −0.007) for CFI and +0.025 SD/year (−0.005 to +0.054) for Cogstate. Restricting first exposure to within 90 days of registration yielded estimates of −0.038 SD/year (−0.058 to −0.009) and +0.021 SD/year (−0.004 to +0.058), respectively (Supplementary Figure S4C; Supplementary Table S4B). All 500 requested rematching-bootstrap replicates were valid for each outcome under both specifications.

Using a narrower calendar matching window of ±90 days produced reference slopes of −0.024 SD/year (95% cluster-robust CI, −0.045 to −0.003) for CFI and +0.020 SD/year (−0.004 to +0.045) for Cogstate. These confidence intervals were obtained using a different procedure and were not compared directly with the rematching-bootstrap intervals as measures of equivalent uncertainty. Across 13 measured entrant-composition variables, the maximum absolute early-versus-later SMD was 0.134 for CFI and 0.137 for Cogstate. These measured differences were modest, although they do not exclude unmeasured differences between enrollment periods.

**S6. Retest-model specifications, estimator differences, and predictive performance**

The complete specification grid included 60 fitted models across the two outcomes, five retest representations, two elapsed-time functions, and three estimators (Supplementary Figure S3; Supplementary Table S3). The marginal and mixed-effects models included 9,611 CFI participants contributing 36,626 assessment records within 36 months and 5,898 Cogstate participants contributing 27,775 records. Participant fixed-effects models used the restricted subsets with repeated complete observations: 5,052 CFI participants and 3,646 Cogstate participants.

Under linear elapsed time, the marginal CFI coefficients in favorable baseline-SD units were +0.080 SD/year with categorical assessment-number adjustment, −0.117 with a post-first indicator plus linear log assessment number, −0.129 with expanded assessment-number categories, −0.216 with exact assessment-number indicators through occasion 10, and −0.227 with the spline-log representation. The corresponding Cogstate coefficients were +0.265, +0.033, +0.057, −0.033, and −0.049 SD/year, respectively. Mixed-effects and fixed-effects models also showed substantial variation across retest specifications (Supplementary Table S3).

The differences remained evident when elapsed time was modeled flexibly. Under a natural-spline time function, CFI marginal annualized 0–36-month contrasts ranged from +0.132 SD/year with categorical assessment-number adjustment to −0.240 SD/year with the post-first plus linear log assessment-number specification. The corresponding Cogstate contrasts ranged from +0.224 SD/year with categorical adjustment to −0.086 SD/year with exact assessment-number indicators through occasion 10. Under the post-first plus spline-log retest specification, the annualized contrasts were −0.166 SD/year for CFI and −0.076 SD/year for Cogstate. Model-implied assessment-number contrasts also differed by retest representation (Supplementary Figure S3C,D).

Within the linear-time models, AIC favored the spline-log retest specification among the evaluated marginal and mixed-effects models. Compared with that specification, the categorical 1/2/3/4+ representation had delta AIC values of 111.9 for CFI and 91.5 for Cogstate in marginal regression, and 76.1 and 99.0, respectively, in mixed-effects modeling. In contrast, participant-clustered cross-validated prediction errors differed little among marginal specifications: the maximum excess RMSE was approximately 0.125% for CFI and 0.377% for Cogstate across the evaluated time functions. Across linear-time marginal models, constraining the elapsed-time coefficient to the first-exposure reference slope changed cross-validated RMSE by −0.020% to +1.121%, depending on outcome and retest specification (Supplementary Table S3). These results describe prediction sensitivity to the imposed constraint rather than establishing the validity of the reference slope.

Collectively, the expanded specification analyses showed that modest differences in predictive performance could coexist with substantially different estimated elapsed-time coefficients.

In time-origin sensitivity analyses, using elapsed time since registration rather than since the first valid assessment changed the categorical-retest marginal coefficient from +0.080 to +0.067 SD/year for CFI and from +0.265 to +0.183 SD/year for Cogstate. Additional adjustment for the delay between registration and the first valid assessment produced little change in the primary first-assessment-origin time coefficients. Participant fixed-effects coefficients were unchanged by the time-origin transformation. The overall dependence of elapsed-time coefficients on retest representation remained evident across these analyses (Supplementary Table S5).

**S7. Observation-weighting diagnostics and truncation sensitivity**

Observation-weighting diagnostics indicated considerable limitations in the positivity of the estimated assessment-observation process (Supplementary Figure S5; Supplementary Table S4C). The primary risk-interval datasets included 186,081 CFI and 111,299 Cogstate rolling availability intervals. Estimated denominator observation probabilities were below 0.01 for 49.9% and 43.4% of the respective modeled intervals. Under primary 1st–99th percentile weight truncation, effective sample sizes represented only 13.3% of observed weighted CFI assessments and 26.8% of observed weighted Cogstate assessments.

In complete-case outcome models using categorical adjustment for assessment number, the native CFI elapsed-time coefficient was −0.066 points/year (95% CI, −0.097 to −0.035) without weighting and −0.084 points/year (−0.154 to −0.014) with primary IPOW. The corresponding Cogstate coefficients were +0.103 z-score units/year (0.095 to 0.111) and +0.098 (0.085 to 0.111), respectively. These native-unit coefficients should not be compared numerically with the favorable standardized estimates in Supplementary Table S3.

More aggressive weight truncation substantially increased effective sample size while altering the CFI coefficient. With truncation at the 2.5th–97.5th and 5th–95th percentiles, the estimated CFI coefficients were −0.063 and −0.059 points/year, respectively, compared with −0.084 under the primary truncation. The corresponding effective sample-size fractions increased from 13.0% to 60.6% and 70.0% in the complete-case outcome-model sample. For Cogstate, the weighted coefficients remained close to +0.10 z-score units/year, while effective sample-size fractions increased from 26.3% to 66.8% and 77.0%.

Changing the rolling risk-interval width produced primary categorical-model IPOW estimates of −0.091, −0.084, and −0.090 CFI points/year for 60-, 90-, and 120-day intervals, respectively. Corresponding Cogstate estimates were +0.097, +0.098, and +0.101 z-score units/year. Analyses using more flexible observation-model functions gave similar point estimates, but very low estimated observation probabilities remained common.

**S8. Joint sensitivity to retest representation and selective follow-up**

Weighting did not eliminate dependence on how cumulative assessment exposure was modeled (Supplementary Figure S5; Supplementary Table S4C). For CFI, the primary IPOW coefficient was −0.084 points/year under categorical assessment-number adjustment, compared with +0.107 points/year using a post-first indicator plus linear log assessment number and +0.199 points/year using a post-first indicator plus spline-log assessment number. The corresponding Cogstate coefficients were +0.098, +0.017, and +0.013 z-score units/year.

Sensitivity analyses among participants with at least 36 months of potential follow-up also produced coefficients that varied by retest specification. The common-window samples included 8,831 CFI participants and 5,410 Cogstate participants. In several CFI models, confidence intervals were wide, and the results did not identify a single stable estimate of elapsed-time-related change.

These analyses reinforce the descriptive finding that observed assessment history and subsequent observation were interrelated. Given the prevalence of very low estimated observation probabilities, the sensitivity of effective sample size to truncation, and the variation across retest specifications, the IPOW analyses could not establish that informative observation had been adequately corrected or that the remaining elapsed-time coefficient represented underlying biological cognitive change.

**Supplementary Figures**

**Figure S1. Participant-driven assessment timing and the relationship between elapsed time and cumulative assessment number**

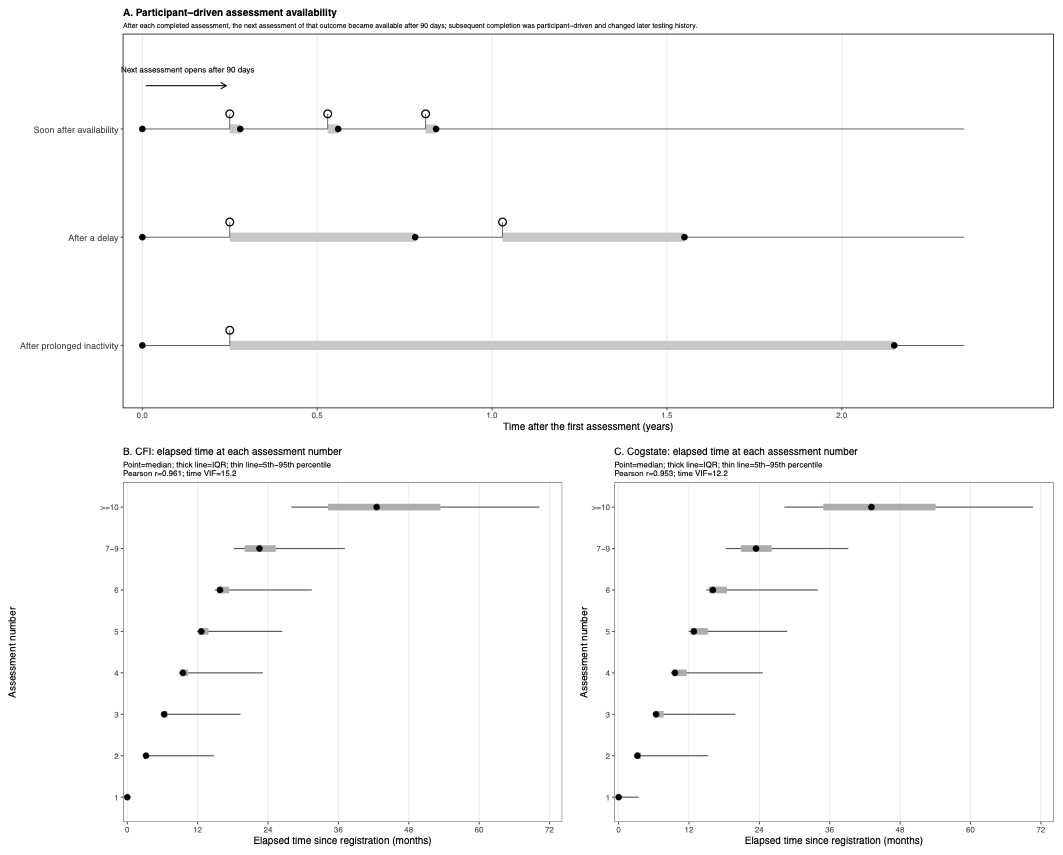

**(A)** Illustrative assessment histories showing how assessments become available 90 days after completion of the preceding assessment of the same instrument. Subsequent completion may occur soon after availability, following a delay, or after prolonged inactivity. These histories are schematic and do not depict individual study participants.

**(B, C)** Distributions of elapsed time since registration by cumulative assessment number for CFI (B) and Cogstate (C). Points indicate medians, thick horizontal intervals indicate interquartile ranges, and thin intervals indicate the 5th–95th percentile ranges. Assessment occasions are grouped at higher cumulative numbers for presentation. Pearson correlation coefficients and variance inflation factors summarize the close relationship between elapsed time and assessment number in the analyzed data.

**Abbreviations:** CFI, Cognitive Function Instrument; IQR, interquartile range; VIF, variance inflation factor.

**Figure S2. Prolonged inactivity, reactivation, and sensitivity to operational definitions**

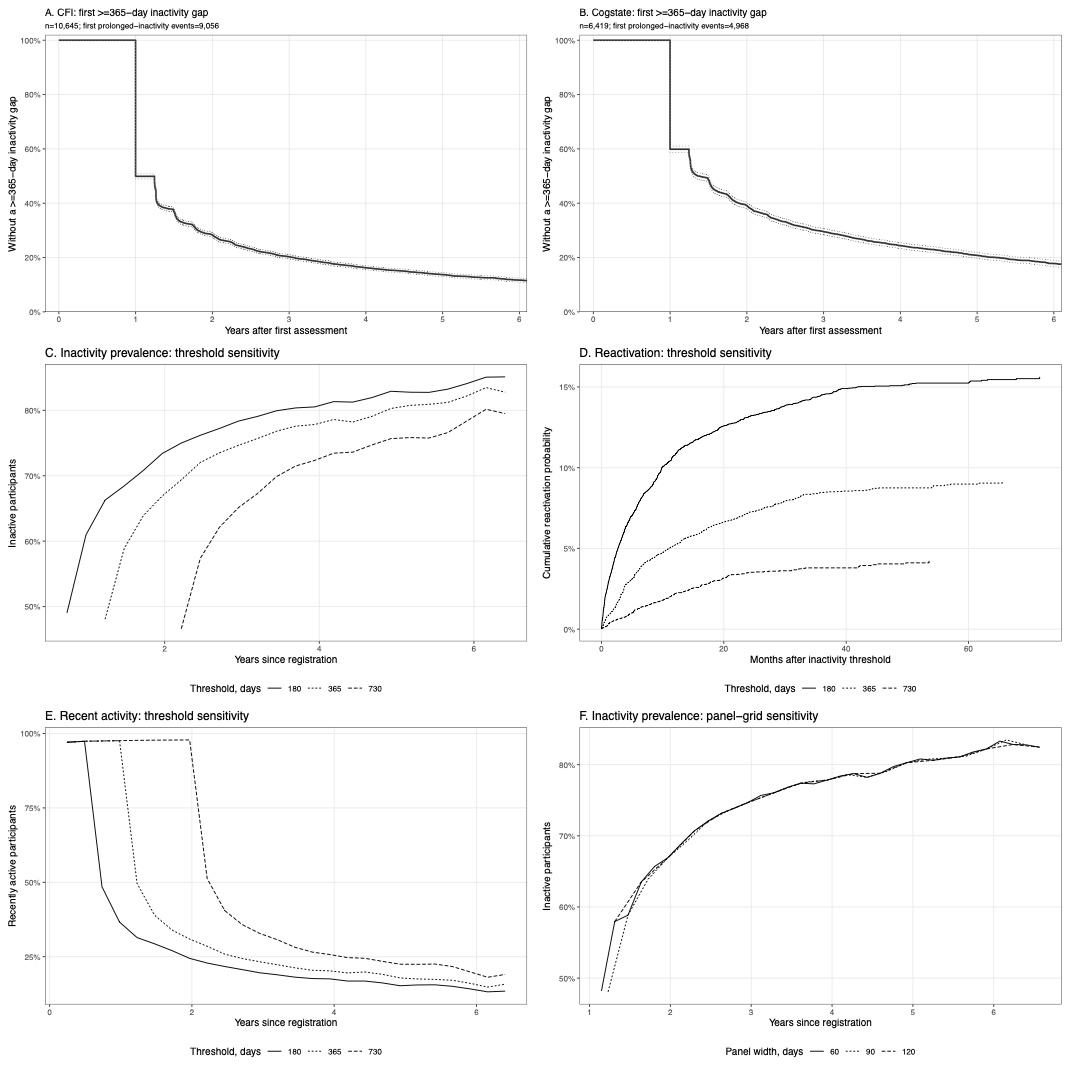

**(A, B)** Kaplan–Meier estimates of the probability of remaining without a first assessment gap of at least 365 consecutive days after the first observed CFI (A) or Cogstate (B) assessment. Participants without a qualifying gap were censored at the administrative observation cutoff.

**(C)** Proportion of participants classified as inactive over time since registration using alternative inactivity thresholds of 180, 365, and 730 days.

**(D)** Kaplan–Meier cumulative reactivation probabilities following the first qualifying inactivity episode under each threshold definition.

**(E)** Proportion classified as recently active over time since registration, using the corresponding activity thresholds.

**(F)** Sensitivity of estimated inactivity prevalence to registration-anchored panel widths of 60, 90, and 120 days, retaining the 365-day inactivity definition.

Time-dependent proportions were evaluated among participants with sufficient observation opportunity. The main analyses used 90-day panels and a 365-day inactivity threshold. Inactivity represents an operational absence of assessment activity rather than confirmed study withdrawal.

**Abbreviations:** CFI, Cognitive Function Instrument; KM, Kaplan–Meier.

**Figure S3. Full comparison of repeated-assessment representations and model-implied score contrasts**

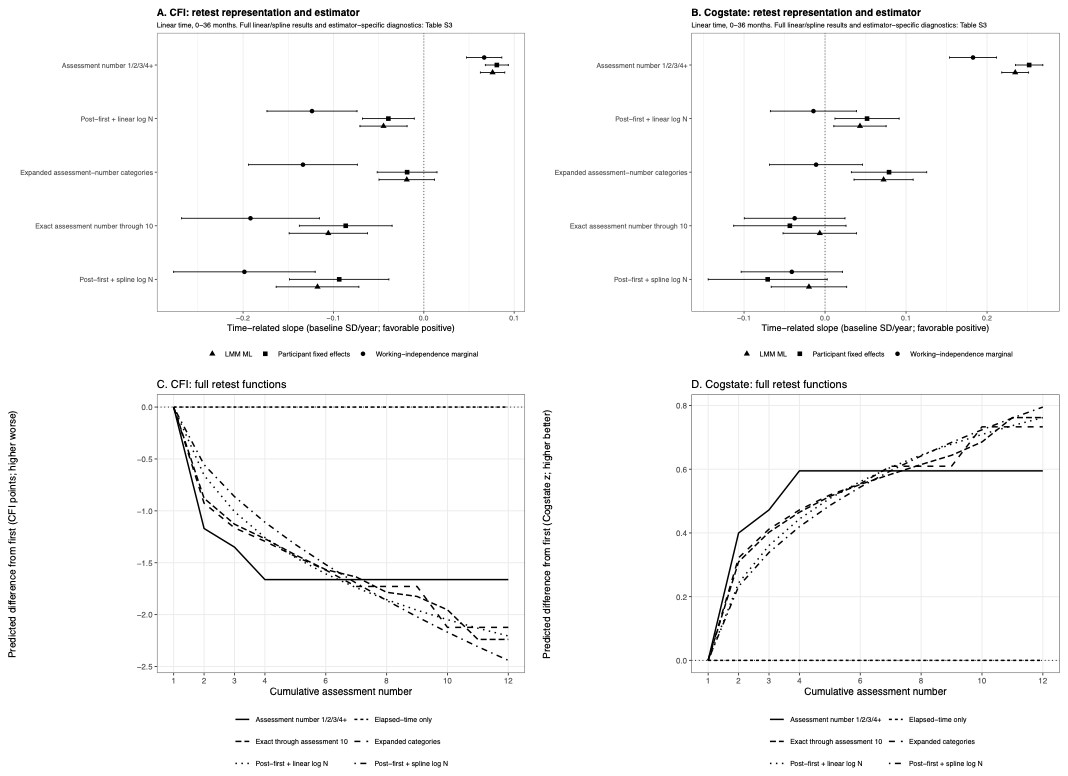

**(A, B)** Estimated annual elapsed-time coefficients for CFI (A) and Cogstate (B) across five representations of repeated-assessment exposure: categorical assessment number (1/2/3/4+), post-first-assessment indicator plus linear log assessment number, expanded assessment-number categories, exact assessment number through the tenth occasion, and post-first-assessment indicator plus spline log assessment number. Models were fitted using working-independence marginal regression, maximum-likelihood linear mixed-effects models, and participant fixed-effects estimation. Linear elapsed time was evaluated over months 0–36 after the first valid assessment of the respective instrument. Points indicate estimates and horizontal intervals represent 95% confidence intervals. Positive values indicate favorable change in baseline-standard-deviation units.

**(C, D)** Model-implied contrasts associated with cumulative assessment number for CFI (C) and Cogstate (D), relative to the first assessment, evaluated at common elapsed-time and covariate values. These illustrative predictions are expressed in the original analytical outcome units: CFI points (higher indicates worse self-reported difficulties) and Cogstate composite z-score units (higher indicates favorable performance under the analytic scoring convention). The elapsed-time-only specification has no assessment-number term and therefore provides a zero reference contrast. The curves represent conditional model predictions rather than observed individual trajectories or causal practice effects.

The complete linear- and spline-time specification grid, including estimator-specific model-fit and prediction diagnostics, is reported in Supplementary Table S3.

**Abbreviations:** CFI, Cognitive Function Instrument; CI, confidence interval; FE, fixed effects; LMM, linear mixed-effects model; SD, standard deviation.

**Figure S4. Contemporaneous first-exposure matching design, matching support, and sensitivity to the timing of first assessments**

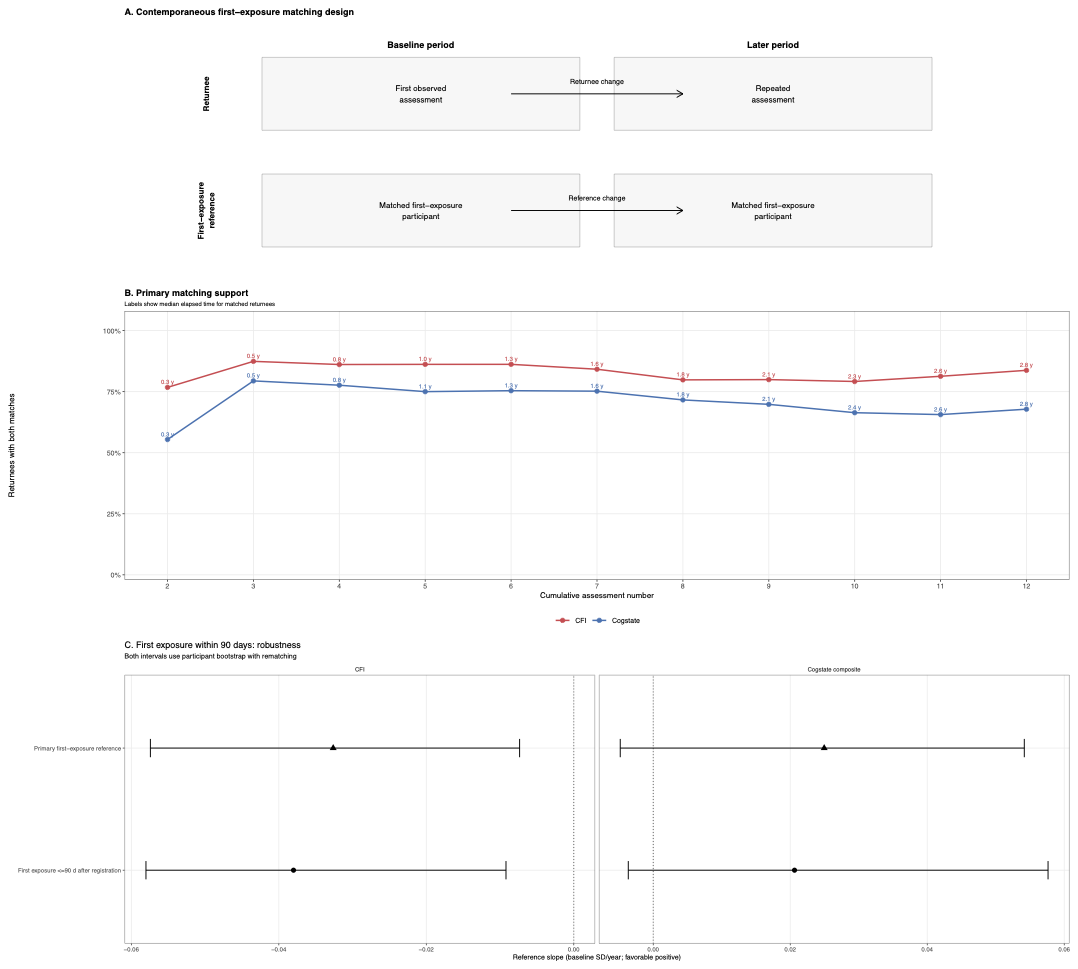

**(A)** Schematic representation of the contemporaneous first-exposure comparison. Each returnee contributes an initial and a repeated assessment, from which within-person change is calculated. Two distinct participants completing their first observed J-TRC assessment are separately matched to the returnee's initial and later assessment dates. Their between-participant score difference constitutes the first-exposure reference contrast. Matching required the same sex, calendar dates within ±180 days, attained ages within ±2 years, and educational attainment within ±2 years.

**(B)** Proportion of eligible returnees successfully matched at both time points, by cumulative assessment number, separately for CFI and Cogstate. Text annotations give the median elapsed time between first and repeated assessments among matched returnees.

**(C)** Participant-equal first-exposure reference slopes under the primary matching specification and a sensitivity analysis restricting first-observed J-TRC assessments to within 90 days after registration. Points represent annual slopes in favorable baseline-standard-deviation units, and horizontal intervals represent 95% percentile confidence intervals from 500 participant-level bootstrap replicates with repeated matching. The vertical dotted line indicates zero.

The reference contrasts are not within-person trajectories of the comparator participants and do not represent biological aging or a pure practice effect. Numerical estimates and other matching sensitivities are provided in Supplementary Tables S4A and S4B.

**Abbreviations:** CFI, Cognitive Function Instrument; CI, confidence interval; J-TRC, Japanese Trial-Ready Cohort; SD, standard deviation.

**Figure S5. Sensitivity of time-associated coefficients to inverse-probability-of-observation weighting and retest adjustment**

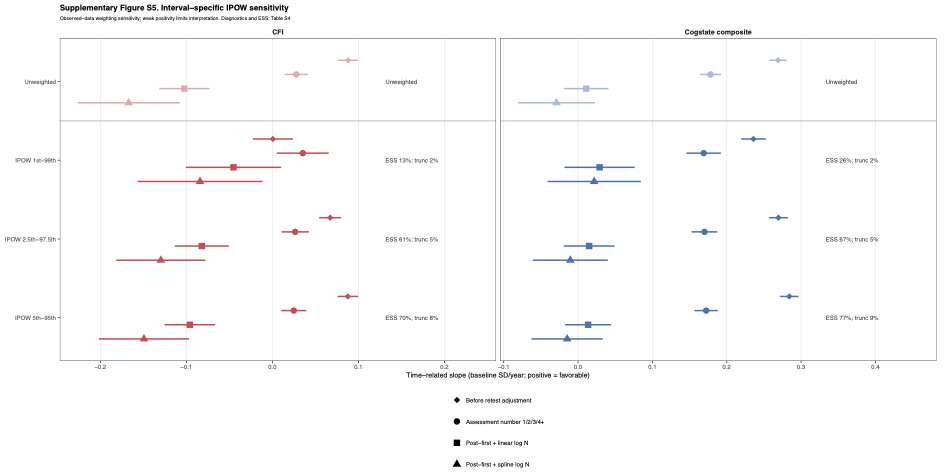

Observed-data estimates of time-associated coefficients for CFI and Cogstate under unweighted analyses and inverse-probability-of-observation weighting (IPOW). Weights were estimated from outcome-specific rolling assessment-availability intervals and truncated at the 1st–99th, 2.5th–97.5th, or 5th–95th percentiles. Within each weighting specification, estimates are presented before explicit retest adjustment (diamonds) and after adjustment using categorical assessment number (circles), post-first-assessment indicator plus linear log assessment number (squares), or post-first-assessment indicator plus spline log assessment number (triangles).

Points represent estimates, and horizontal intervals indicate 95% confidence intervals. Coefficients are expressed in favorable baseline-standard-deviation units per year, with positive values indicating favorable change. Annotations indicate effective sample size as a percentage of observed weighted assessments and the percentage of observations with truncated weights. Supplementary Table S4C provides the corresponding observation-model diagnostics and additional sensitivity analyses; its native-unit coefficients should not be compared numerically with the standardized coefficients displayed here.

Low estimated observation probabilities and reduced effective sample sizes indicate limited positivity and restrict the interpretation of IPOW estimates as corrections for informative observation.

**Abbreviations:** CFI, Cognitive Function Instrument; CI, confidence interval; ESS, effective sample size; IPOW, inverse probability of observation weighting; SD, standard deviation.
