## Supplementary material for "Longitudinal Participation and Cognitive Score Interpretation in the J-TRC Webstudy": Table 1

Tables

**Table 1. Cohort characteristics and longitudinal data contribution in the J-TRC Webstudy**

**Legend and footnotes**

| **Section** | **Measure** | **Value** |
| --- | --- | --- |
| **A. Denominators and longitudinal data contribution** | Registered accounts by inclusion cutoff | 14,912 |
|  | Accounts with explicit affirmative consent | 12,068 |
|  | Consent accounts with eligibility fields ascertainable | 11,901/12,068 (98.6%) |
|  | Eligible consented cohort | 10,863 |
|  | Fully registered eligible subset | 10,347/10,863 (95.2%) |
|  | Completion of at least one valid assessment | 10,645/10,863 (98.0%) |
|  | Completion of at least one valid CFI assessment | 10,645/10,863 (98.0%) |
|  | Valid assessments on at least 2 distinct days | 5,792/10,863 (53.3%) |
|  | Completion of at least one valid Cogstate assessment | 6,419/10,863 (59.1%) |
|  | Contribution at or beyond 12 months | 3,611/10,863 (33.2%) |
|  | Contribution at or beyond 36 months | 1,936/9,977 (19.4%) |
|  | Contribution at or beyond 60 months | 651/4,602 (14.1%) |
|  | Assessment during the final 365 days of observation | 1,825/10,863 (16.8%) |
| **B. Participant characteristics across contribution stages** | Eligible consented cohort | n=10,863; age 62.5 (8.4); female 5,927/10,858 (54.6%); education 15.2 (2.0) |
|  | Participants completing a valid CFI assessment | n=10,645; age 62.5 (8.4); female 5,808/10,640 (54.6%); education 15.2 (2.0) |
|  | Participants completing a valid Cogstate assessment | n=6,419; age 62.6 (8.2); female 3,220/6,419 (50.2%); education 15.4 (1.9) |
|  | Participants with repeated activity | n=5,792; age 62.2 (8.1); female 3,159/5,792 (54.5%); education 15.3 (1.9) |

Panel A summarizes registry-level denominators, completion of initial valid assessments, repeated assessment activity, and longitudinal data contribution. Registered accounts were restricted to those registered by March 31, 2025, with assessment data available through May 31, 2026. The eligible consented cohort comprised participants aged 50–85 years with explicit affirmative consent, ascertainable eligibility information, and no self-reported diagnosis of Alzheimer's disease. Initial assessment completion was defined as at least one valid CFI or Cogstate assessment, and repeated activity was defined as valid assessment activity on at least two distinct calendar days, regardless of instrument. Longitudinal contribution at or beyond 12, 36, and 60 months required at least one assessment at or beyond the respective time threshold; denominators were restricted to participants with sufficient observation opportunity.

Panel B presents characteristics of the eligible consented cohort and selected assessment-contributing populations. Age and educational attainment are reported as mean (standard deviation), and female sex as n/N (%), using available observations for each characteristic. More detailed cohort composition, registration-year distributions, and comparisons across participation stages are provided in Supplementary Table S1.

**Abbreviations:** AD, Alzheimer's disease; CFI, Cognitive Function Instrument; J-TRC, Japanese Trial-Ready Cohort; n, number; N, denominator; SD, standard deviation.
