## Supplementary Tables for "Longitudinal Participation and Cognitive Score Interpretation in the J-TRC Webstudy"

### Supplementary Table S1. Cohort characteristics, scoring/QC, and analysis populations

| Section | Outcome | Analysis | Estimate or result | Sample | Additional information |
|---|---|---|---|---|---|
| **Cohort construction** | | Registered accounts by inclusion cutoff | 14,912 | | Registration by March 31, 2025. |
| **Cohort construction** | | Accounts with explicit affirmative consent | 12,068 | | Defined only by an affirmative consent response; workflow stage alone was not counted as consent. |
| **Cohort construction** | | Consent accounts with eligibility fields ascertainable | 11,901/12,068 (98.6%) | | Age and self-reported AD status were both observed; valid assessment completion and final workflow completion were not required. |
| **Cohort construction** | | Eligible consented cohort | 10,863 | | Affirmative consent, ascertainable age and AD status, no self-reported AD, age 50–85 years, and registration by the inclusion cutoff. |
| **Cohort construction** | | Fully registered eligible subset | 10,347/10,863 (95.2%) | | Secondary subset additionally reaching the final information-registration workflow stage; not used as the primary observability denominator. |
| **Cohort construction** | | Completion of at least one valid assessment | 10,645/10,863 (98.0%) | | At least one valid CFI or Cogstate activity day. |
| **Cohort construction** | | Valid assessments on at least 2 distinct days | 5,792/10,863 (53.3%) | | At least two distinct activity days. |
| **Cohort construction** | | Completion of at least one valid Cogstate assessment | 6,419/10,863 (59.1%) | | At least one Cogstate activity day. |
| **Cohort construction** | | Contribution at or beyond 12 months | 3,611/10,863 (33.2%) | | Denominator restricted to sufficient follow-up opportunity. |
| **Cohort construction** | | Consent accounts with eligibility fields not ascertainable | 167/12,068 (1.4%) | | Age and/or self-reported AD status was unavailable; these accounts were not classified as eligible or ineligible. |
| **Analytic data availability** | | Valid CFI assessment days in analytic outcome dataset | 51,384 | | Latest valid CFI retained per participant-day. |
| **Analytic data availability** | | Valid Cogstate sessions in analytic outcome dataset | 40,058 | | Exact timestamp duplicates removed; primary composite required at least two domains. |
| **Analytic data availability** | | CFI same-sample 90-day analysis records | 9,611 participants; 46,716 90-day analysis records | | Identical rows across the five retest specifications and the time-only benchmark. |
| **Analytic data availability** | | Cogstate same-sample 90-day analysis records | 5,898 participants; 36,614 90-day analysis records | | Identical rows across the five retest specifications and the time-only benchmark. |
| **Selection across contribution stages** | | Completion of at least one valid CFI assessment: Age at registration | 0.000 | | Standardized mean difference comparing participants inside versus outside the stage. |
| **Selection across contribution stages** | | Completion of at least one valid CFI assessment: Female sex | -0.000 | | Standardized mean difference comparing participants inside versus outside the stage. |
| **Selection across contribution stages** | | Completion of at least one valid CFI assessment: Education | 0.205 | | Standardized mean difference comparing participants inside versus outside the stage. |
| **Selection across contribution stages** | | Completion of at least one valid CFI assessment: Registration year 2019 | 0.134 | | Standardized mean difference comparing participants inside versus outside the stage. |
| **Selection across contribution stages** | | Completion of at least one valid CFI assessment: Registration year 2020 | -0.008 | | Standardized mean difference comparing participants inside versus outside the stage. |
| **Selection across contribution stages** | | Completion of at least one valid CFI assessment: Registration year 2021 | -0.074 | | Standardized mean difference comparing participants inside versus outside the stage. |
| **Selection across contribution stages** | | Completion of at least one valid CFI assessment: Registration year 2022 | 0.056 | | Standardized mean difference comparing participants inside versus outside the stage. |
| **Selection across contribution stages** | | Completion of at least one valid CFI assessment: Registration year 2023 | -0.128 | | Standardized mean difference comparing participants inside versus outside the stage. |
| **Selection across contribution stages** | | Completion of at least one valid CFI assessment: Registration year 2024 | -0.002 | | Standardized mean difference comparing participants inside versus outside the stage. |
| **Selection across contribution stages** | | Completion of at least one valid CFI assessment: Registration year 2025 | -0.072 | | Standardized mean difference comparing participants inside versus outside the stage. |
| **Selection across contribution stages** | | Completion of at least one valid Cogstate assessment: Age at registration | 0.038 | | Standardized mean difference comparing participants inside versus outside the stage. |
| **Selection across contribution stages** | | Completion of at least one valid Cogstate assessment: Female sex | -0.219 | | Standardized mean difference comparing participants inside versus outside the stage. |
| **Selection across contribution stages** | | Completion of at least one valid Cogstate assessment: Education | 0.242 | | Standardized mean difference comparing participants inside versus outside the stage. |
| **Selection across contribution stages** | | Completion of at least one valid Cogstate assessment: Registration year 2019 | 0.176 | | Standardized mean difference comparing participants inside versus outside the stage. |
| **Selection across contribution stages** | | Completion of at least one valid Cogstate assessment: Registration year 2020 | -0.002 | | Standardized mean difference comparing participants inside versus outside the stage. |
| **Selection across contribution stages** | | Completion of at least one valid Cogstate assessment: Registration year 2021 | -0.111 | | Standardized mean difference comparing participants inside versus outside the stage. |
| **Selection across contribution stages** | | Completion of at least one valid Cogstate assessment: Registration year 2022 | 0.008 | | Standardized mean difference comparing participants inside versus outside the stage. |
| **Selection across contribution stages** | | Completion of at least one valid Cogstate assessment: Registration year 2023 | -0.102 | | Standardized mean difference comparing participants inside versus outside the stage. |
| **Selection across contribution stages** | | Completion of at least one valid Cogstate assessment: Registration year 2024 | 0.010 | | Standardized mean difference comparing participants inside versus outside the stage. |
| **Selection across contribution stages** | | Completion of at least one valid Cogstate assessment: Registration year 2025 | -0.028 | | Standardized mean difference comparing participants inside versus outside the stage. |
| **Selection across contribution stages** | | Repeated activity: Age at registration | -0.070 | | Standardized mean difference comparing participants inside versus outside the stage. |
| **Selection across contribution stages** | | Repeated activity: Female sex | -0.002 | | Standardized mean difference comparing participants inside versus outside the stage. |
| **Selection across contribution stages** | | Repeated activity: Education | 0.110 | | Standardized mean difference comparing participants inside versus outside the stage. |
| **Selection across contribution stages** | | Repeated activity: Registration year 2019 | 0.091 | | Standardized mean difference comparing participants inside versus outside the stage. |
| **Selection across contribution stages** | | Repeated activity: Registration year 2020 | -0.031 | | Standardized mean difference comparing participants inside versus outside the stage. |
| **Selection across contribution stages** | | Repeated activity: Registration year 2021 | -0.070 | | Standardized mean difference comparing participants inside versus outside the stage. |
| **Selection across contribution stages** | | Repeated activity: Registration year 2022 | 0.069 | | Standardized mean difference comparing participants inside versus outside the stage. |
| **Selection across contribution stages** | | Repeated activity: Registration year 2023 | -0.089 | | Standardized mean difference comparing participants inside versus outside the stage. |
| **Selection across contribution stages** | | Repeated activity: Registration year 2024 | -0.021 | | Standardized mean difference comparing participants inside versus outside the stage. |
| **Selection across contribution stages** | | Repeated activity: Registration year 2025 | -0.053 | | Standardized mean difference comparing participants inside versus outside the stage. |
| **Selection across contribution stages** | | Active within 365 days: Age at registration | 0.113 | | Standardized mean difference comparing participants inside versus outside the stage. |
| **Selection across contribution stages** | | Active within 365 days: Female sex | -0.085 | | Standardized mean difference comparing participants inside versus outside the stage. |
| **Selection across contribution stages** | | Active within 365 days: Education | 0.129 | | Standardized mean difference comparing participants inside versus outside the stage. |
| **Selection across contribution stages** | | Active within 365 days: Registration year 2019 | -0.040 | | Standardized mean difference comparing participants inside versus outside the stage. |
| **Selection across contribution stages** | | Active within 365 days: Registration year 2020 | -0.072 | | Standardized mean difference comparing participants inside versus outside the stage. |
| **Selection across contribution stages** | | Active within 365 days: Registration year 2021 | -0.101 | | Standardized mean difference comparing participants inside versus outside the stage. |
| **Selection across contribution stages** | | Active within 365 days: Registration year 2022 | 0.045 | | Standardized mean difference comparing participants inside versus outside the stage. |
| **Selection across contribution stages** | | Active within 365 days: Registration year 2023 | 0.012 | | Standardized mean difference comparing participants inside versus outside the stage. |
| **Selection across contribution stages** | | Active within 365 days: Registration year 2024 | 0.176 | | Standardized mean difference comparing participants inside versus outside the stage. |
| **Selection across contribution stages** | | Active within 365 days: Registration year 2025 | 0.098 | | Standardized mean difference comparing participants inside versus outside the stage. |
| **Selection across contribution stages** | | Contribution at or beyond 36 months: Age at registration | 0.082 | | Standardized mean difference comparing participants inside versus outside the stage. |
| **Selection across contribution stages** | | Contribution at or beyond 36 months: Female sex | -0.078 | | Standardized mean difference comparing participants inside versus outside the stage. |
| **Selection across contribution stages** | | Contribution at or beyond 36 months: Education | 0.115 | | Standardized mean difference comparing participants inside versus outside the stage. |
| **Selection across contribution stages** | | Contribution at or beyond 36 months: Registration year 2019 | 0.205 | | Standardized mean difference comparing participants inside versus outside the stage. |
| **Selection across contribution stages** | | Contribution at or beyond 36 months: Registration year 2020 | 0.065 | | Standardized mean difference comparing participants inside versus outside the stage. |
| **Selection across contribution stages** | | Contribution at or beyond 36 months: Registration year 2021 | -0.047 | | Standardized mean difference comparing participants inside versus outside the stage. |
| **Selection across contribution stages** | | Contribution at or beyond 36 months: Registration year 2022 | -0.095 | | Standardized mean difference comparing participants inside versus outside the stage. |
| **Selection across contribution stages** | | Contribution at or beyond 36 months: Registration year 2023 | -0.226 | | Standardized mean difference comparing participants inside versus outside the stage. |
| **Selection across contribution stages** | | Contribution at or beyond 36 months: Registration year 2024 | NA (no 36-month observation opportunity) | | Standardized mean difference comparing participants inside versus outside the stage. |
| **Selection across contribution stages** | | Contribution at or beyond 36 months: Registration year 2025 | NA (no 36-month observation opportunity) | | Standardized mean difference comparing participants inside versus outside the stage. |
| **Contribution-stage characteristics and registration years** | | Eligible consented cohort | Age 62.5 (8.4); female 5,927/10,858 (54.6%); education 15.2 (2.0) | 10,863 participants | Registration years among nonmissing values: 2019: 2,021 (18.6%); 2020: 1,319 (12.1%); 2021: 1,793 (16.5%); 2022: 4,326 (39.8%); 2023: 886 (8.2%); 2024: 445 (4.1%); 2025: 73 (0.7%); missing age/sex/education/registration year = 0/5/1074/0 |
| **Contribution-stage characteristics and registration years** | | Participants completing a valid CFI assessment | Age 62.5 (8.4); female 5,808/10,640 (54.6%); education 15.2 (2.0) | 10,645 participants | Registration years among nonmissing values: 2019: 1,991 (18.7%); 2020: 1,292 (12.1%); 2021: 1,751 (16.4%); 2022: 4,245 (39.9%); 2023: 860 (8.1%); 2024: 436 (4.1%); 2025: 70 (0.7%); missing age/sex/education/registration year = 0/5/1030/0 |
| **Contribution-stage characteristics and registration years** | | Participants completing a valid Cogstate assessment | Age 62.6 (8.2); female 3,220/6,419 (50.2%); education 15.4 (1.9) | 6,419 participants | Registration years among nonmissing values: 2019: 1,371 (21.4%); 2020: 778 (12.1%); 2021: 950 (14.8%); 2022: 2,566 (40.0%); 2023: 449 (7.0%); 2024: 268 (4.2%); 2025: 37 (0.6%); missing age/sex/education/registration year = 0/0/521/0 |
| **Contribution-stage characteristics and registration years** | | Repeated activity | Age 62.2 (8.1); female 3,159/5,792 (54.5%); education 15.3 (1.9) | 5,792 participants | Registration years among nonmissing values: 2019: 1,173 (20.3%); 2020: 676 (11.7%); 2021: 886 (15.3%); 2022: 2,398 (41.4%); 2023: 406 (7.0%); 2024: 226 (3.9%); 2025: 27 (0.5%); missing age/sex/education/registration year = 0/0/523/0 |
| **Contribution-stage characteristics and registration years** | | Assessment during the final 365 days of observation | Age 63.3 (7.9); female 932/1,825 (51.1%); education 15.4 (1.9) | 1,825 participants | Registration years among nonmissing values: 2019: 316 (17.3%); 2020: 187 (10.2%); 2021: 246 (13.5%); 2022: 760 (41.6%); 2023: 154 (8.4%); 2024: 135 (7.4%); 2025: 27 (1.5%); missing age/sex/education/registration year = 0/0/154/0 |
| **Contribution-stage characteristics and registration years** | | Contribution at or beyond 36 months | Age 62.9 (7.9); female 996/1,936 (51.4%); education 15.4 (1.9) | 1,936 participants | Registration years among nonmissing values: 2019: 526 (27.2%); 2020: 291 (15.0%); 2021: 320 (16.5%); 2022: 766 (39.6%); 2023: 33 (1.7%); missing age/sex/education/registration year = 0/0/168/0 |
| **Characteristics of 0-36-month analytic populations** | | CFI same-sample analytic participants | n=9,611; age 62.7 (8.4); female 4,982/9,611 (51.8%); education 15.2 (2.0) | | Age and education are mean (SD); female sex is n/N (%). Registration-year distributions and selection contrasts are reported in Supplementary Table S1. |
| **Characteristics of 0-36-month analytic populations** | | Cogstate same-sample analytic participants | n=5,898; age 62.9 (8.2); female 2,809/5,898 (47.6%); education 15.4 (1.9) | | Age and education are mean (SD); female sex is n/N (%). Registration-year distributions and selection contrasts are reported in Supplementary Table S1. |
| **CFI does-not-apply frequency** | | Overall: All valid CFI assessment days | 18,188/51,384 (35.40%) | | Median 0.0; maximum 3 does-not-apply items per session. |
| **CFI does-not-apply frequency** | | Assessment number: 1 | 3,058/10,645 (28.73%) | | Median 0.0; maximum 3 does-not-apply items per session. |
| **CFI does-not-apply frequency** | | Assessment number: 2 | 1,832/5,610 (32.66%) | | Median 0.0; maximum 3 does-not-apply items per session. |
| **CFI does-not-apply frequency** | | Assessment number: 3 | 1,446/4,219 (34.27%) | | Median 0.0; maximum 3 does-not-apply items per session. |
| **CFI does-not-apply frequency** | | Assessment number: 4plus | 11,852/30,910 (38.34%) | | Median 0.0; maximum 3 does-not-apply items per session. |
| **CFI scoring sensitivity** | | All 15 items scorable: Elapsed-time-only model | -0.238 (-0.270 to -0.205) | | 7,467 participants; 30,435 90-day analysis records |
| **CFI scoring sensitivity** | | All 15 items scorable: Categorical assessment number (1/2/3/4+) | -0.084 (-0.124 to -0.045) | | 7,467 participants; 30,435 90-day analysis records |
| **CFI scoring sensitivity** | | All 15 items scorable: Post-first + linear log assessment number | 0.230 (0.146 to 0.315) | | 7,467 participants; 30,435 90-day analysis records |
| **CFI scoring sensitivity** | | All 15 items scorable: Expanded assessment-number categories | 0.097 (0.028 to 0.166) | | 7,467 participants; 30,435 90-day analysis records |
| **CFI scoring sensitivity** | | All 15 items scorable: Exact assessment number through 10 | 0.134 (0.056 to 0.213) | | 7,467 participants; 30,435 90-day analysis records |
| **CFI scoring sensitivity** | | All 15 items scorable: Post-first + spline of log assessment number | 0.391 (0.225 to 0.558) | | 7,467 participants; 30,435 90-day analysis records |
| **CFI scoring sensitivity** | | Prorated to 15 items: Elapsed-time-only model | -0.205 (-0.232 to -0.178) | | 9,611 participants; 46,716 90-day analysis records |
| **CFI scoring sensitivity** | | Prorated to 15 items: Categorical assessment number (1/2/3/4+) | -0.061 (-0.093 to -0.029) | | 9,611 participants; 46,716 90-day analysis records |
| **CFI scoring sensitivity** | | Prorated to 15 items: Post-first + linear log assessment number | 0.253 (0.182 to 0.323) | | 9,611 participants; 46,716 90-day analysis records |
| **CFI scoring sensitivity** | | Prorated to 15 items: Expanded assessment-number categories | 0.121 (0.066 to 0.177) | | 9,611 participants; 46,716 90-day analysis records |
| **CFI scoring sensitivity** | | Prorated to 15 items: Exact assessment number through 10 | 0.159 (0.096 to 0.222) | | 9,611 participants; 46,716 90-day analysis records |
| **CFI scoring sensitivity** | | Prorated to 15 items: Post-first + spline of log assessment number | 0.406 (0.261 to 0.552) | | 9,611 participants; 46,716 90-day analysis records |
| **CFI recorded-versus-recalculated total QC** | | Overall | 51,395/51,395 exact matches (100.0%) | | Valid sessions before participant-day collapse. Median absolute difference 0.000; range 0.000 to 0.000. |
| **CFI recorded-versus-recalculated total QC** | | No does-not-apply responses | 33,204/33,204 exact matches (100.0%) | | Valid sessions before participant-day collapse. Median absolute difference 0.000; range 0.000 to 0.000. |
| **CFI recorded-versus-recalculated total QC** | | At least one does-not-apply response | 18,191/18,191 exact matches (100.0%) | | Valid sessions before participant-day collapse. Median absolute difference 0.000; range 0.000 to 0.000. |
| **Cogstate construction and quality control** | | Primary analyzable Cogstate sessions | 40,058 | | At least two available domains required for the primary composite. |
| **Cogstate construction and quality control** | | Sessions with all four prespecified domains | 40,058 | | Reported within the exported analyzable session dataset. |
| **Cogstate construction and quality control** | | Participant-days with more than one genuine all-domain session | 0 | | Multiple genuine same-day sessions remain distinguishable in the source data. |
| **Baseline covariate missingness before complete-case exclusion** | CFI | Sex | 5/51,310 records (0.01%) | 51,310 candidate assessment records | 5 participants with at least one missing record; denominator includes records before complete-case exclusion |
| **Baseline covariate missingness before complete-case exclusion** | CFI | Educational attainment | 4,590/51,310 records (8.95%) | 51,310 candidate assessment records | 1,030 participants with at least one missing record; denominator includes records before complete-case exclusion |
| **Baseline covariate missingness before complete-case exclusion** | Cogstate composite | Educational attainment | 3,376/39,990 records (8.44%) | 39,990 candidate assessment records | 521 participants with at least one missing record; denominator includes records before complete-case exclusion |
| **Complete-case model participants versus excluded participants** | CFI | Retained in complete-case models | Female 51.8%; mean age 62.7 years; mean education 15.2 years | 9,611 participants | Participant-level descriptive characteristics; means use nonmissing values. |
| **Complete-case model participants versus excluded participants** | CFI | Excluded for missing model covariates | Female 80.3%; mean age 60.5 years; mean education 15.0 years | 1,034 participants | Participant-level descriptive characteristics; means use nonmissing values. |
| **Complete-case model participants versus excluded participants** | Cogstate composite | Excluded for missing model covariates | Female 78.9%; mean age 60.0 years; mean education not available (all values missing) | 521 participants | Mean education cannot be calculated because all values are missing in this group. |
| **Complete-case model participants versus excluded participants** | Cogstate composite | Retained in complete-case models | Female 47.6%; mean age 62.9 years; mean education 15.4 years | 5,898 participants | Participant-level descriptive characteristics; means use nonmissing values. |

**Legend:**

Cohort construction, contribution-stage demographics, registration-year distributions, selection contrasts, and scoring quality checks. Covariate missingness percentages use candidate assessment records before complete-case exclusion; retained/excluded characteristics refer to participants. Other model covariates had no missing values in the candidate records.

**Abbreviations:**

CFI, Cognitive Function Instrument; QC, quality control; SD, standard deviation; SMD, standardized mean difference.

### Supplementary Table S2. Participation, inactivity, reactivation, and next observation

| Section | Outcome | Analysis | Estimate or result | Sample | Additional information |
|---|---|---|---|---|---|
| **Long-term contribution and activity before the observation cutoff** | | Contribution at or beyond 12 months | 3,611/10,863 (33.2%) | | Denominator restricted to sufficient follow-up opportunity. |
| **Long-term contribution and activity before the observation cutoff** | | Contribution at or beyond 36 months | 1,936/9,977 (19.4%) | | Denominator restricted to sufficient follow-up opportunity. |
| **Long-term contribution and activity before the observation cutoff** | | Contribution at or beyond 60 months | 651/4,602 (14.1%) | | Denominator restricted to sufficient follow-up opportunity. |
| **Long-term contribution and activity before the observation cutoff** | | Assessment during the final 365 days of observation | 1,825/10,863 (16.8%) | | At least one valid activity day during the preceding 365 days. |
| **Outcome-specific long-term yield with sufficient follow-up opportunity** | Any assessment | Assessment contribution at or beyond 12 months after registration | 3,611/10,863 (33.2%) | 10,863 participants with sufficient follow-up opportunity | At least one valid assessment at or beyond the stated horizon; not assessment completion at a fixed visit. |
| **Outcome-specific long-term yield with sufficient follow-up opportunity** | Any assessment | Assessment contribution at or beyond 36 months after registration | 1,936/9,977 (19.4%) | 9,977 participants with sufficient follow-up opportunity | At least one valid assessment at or beyond the stated horizon; not assessment completion at a fixed visit. |
| **Outcome-specific long-term yield with sufficient follow-up opportunity** | Any assessment | Assessment contribution at or beyond 60 months after registration | 651/4,602 (14.1%) | 4,602 participants with sufficient follow-up opportunity | At least one valid assessment at or beyond the stated horizon; not assessment completion at a fixed visit. |
| **Outcome-specific long-term yield with sufficient follow-up opportunity** | CFI | Assessment contribution at or beyond 12 months after registration | 3,599/10,863 (33.1%) | 10,863 participants with sufficient follow-up opportunity | At least one valid assessment at or beyond the stated horizon; not assessment completion at a fixed visit. |
| **Outcome-specific long-term yield with sufficient follow-up opportunity** | CFI | Assessment contribution at or beyond 36 months after registration | 1,927/9,977 (19.3%) | 9,977 participants with sufficient follow-up opportunity | At least one valid assessment at or beyond the stated horizon; not assessment completion at a fixed visit. |
| **Outcome-specific long-term yield with sufficient follow-up opportunity** | CFI | Assessment contribution at or beyond 60 months after registration | 644/4,602 (14.0%) | 4,602 participants with sufficient follow-up opportunity | At least one valid assessment at or beyond the stated horizon; not assessment completion at a fixed visit. |
| **Outcome-specific long-term yield with sufficient follow-up opportunity** | Cogstate | Assessment contribution at or beyond 12 months after registration | 2,913/10,863 (26.8%) | 10,863 participants with sufficient follow-up opportunity | At least one valid assessment at or beyond the stated horizon; not assessment completion at a fixed visit. |
| **Outcome-specific long-term yield with sufficient follow-up opportunity** | Cogstate | Assessment contribution at or beyond 36 months after registration | 1,689/9,977 (16.9%) | 9,977 participants with sufficient follow-up opportunity | At least one valid assessment at or beyond the stated horizon; not assessment completion at a fixed visit. |
| **Outcome-specific long-term yield with sufficient follow-up opportunity** | Cogstate | Assessment contribution at or beyond 60 months after registration | 595/4,602 (12.9%) | 4,602 participants with sufficient follow-up opportunity | At least one valid assessment at or beyond the stated horizon; not assessment completion at a fixed visit. |
| **Primary 90-day panel and 365-day inactivity definition** | | Year 0.24: Recently active | 10,544/10,863 (97.1%) | | Eligible consented cohort; valid assessment completion was not required. |
| **Primary 90-day panel and 365-day inactivity definition** | | Year 0.24: No previous valid assessment | 319/10,863 (2.9%) | | Eligible consented cohort; valid assessment completion was not required. |
| **Primary 90-day panel and 365-day inactivity definition** | | Year 1.23: Recently active | 5,375/10,833 (49.6%) | | Eligible consented cohort; valid assessment completion was not required. |
| **Primary 90-day panel and 365-day inactivity definition** | | Year 1.23: Inactive ≥365 days | 5,208/10,833 (48.1%) | | Eligible consented cohort; valid assessment completion was not required. |
| **Primary 90-day panel and 365-day inactivity definition** | | Year 1.23: No previous valid assessment | 250/10,833 (2.3%) | | Eligible consented cohort; valid assessment completion was not required. |
| **Primary 90-day panel and 365-day inactivity definition** | | Year 2.21: Recently active | 2,969/10,418 (28.5%) | | Eligible consented cohort; valid assessment completion was not required. |
| **Primary 90-day panel and 365-day inactivity definition** | | Year 2.21: Inactive ≥365 days | 7,226/10,418 (69.4%) | | Eligible consented cohort; valid assessment completion was not required. |
| **Primary 90-day panel and 365-day inactivity definition** | | Year 2.21: No previous valid assessment | 223/10,418 (2.1%) | | Eligible consented cohort; valid assessment completion was not required. |
| **Primary 90-day panel and 365-day inactivity definition** | | Year 3.20: Inactive ≥365 days | 7,279/9,617 (75.7%) | | Eligible consented cohort; valid assessment completion was not required. |
| **Primary 90-day panel and 365-day inactivity definition** | | Year 3.20: Recently active | 2,144/9,617 (22.3%) | | Eligible consented cohort; valid assessment completion was not required. |
| **Primary 90-day panel and 365-day inactivity definition** | | Year 3.20: No previous valid assessment | 194/9,617 (2.0%) | | Eligible consented cohort; valid assessment completion was not required. |
| **Primary 90-day panel and 365-day inactivity definition** | | Year 5.17: Inactive ≥365 days | 3,412/4,225 (80.8%) | | Eligible consented cohort; valid assessment completion was not required. |
| **Primary 90-day panel and 365-day inactivity definition** | | Year 5.17: Recently active | 740/4,225 (17.5%) | | Eligible consented cohort; valid assessment completion was not required. |
| **Primary 90-day panel and 365-day inactivity definition** | | Year 5.17: No previous valid assessment | 73/4,225 (1.7%) | | Eligible consented cohort; valid assessment completion was not required. |
| **Primary 90-day panel and 365-day inactivity definition** | | Year 6.16: Inactive ≥365 days | 2,348/2,813 (83.5%) | | Eligible consented cohort; valid assessment completion was not required. |
| **Primary 90-day panel and 365-day inactivity definition** | | Year 6.16: Recently active | 416/2,813 (14.8%) | | Eligible consented cohort; valid assessment completion was not required. |
| **Primary 90-day panel and 365-day inactivity definition** | | Year 6.16: No previous valid assessment | 49/2,813 (1.7%) | | Eligible consented cohort; valid assessment completion was not required. |
| **Time to reactivation after first 365-day inactivity episode** | | Within 183 days | 3.6% (95% CI 3.3% to 4.0%) | | 8,456 participants at risk. |
| **Time to reactivation after first 365-day inactivity episode** | | Within 365 days | 5.2% (95% CI 4.8% to 5.7%) | | 7,983 participants at risk. |
| **Time to reactivation after first 365-day inactivity episode** | | Within 730 days | 7.2% (95% CI 6.7% to 7.8%) | | 6,896 participants at risk. |
| **Alternative inactivity thresholds** | | 180-day inactivity; reactivation within 183 days | 7.8% (95% CI 7.2% to 8.3%) | | 8,561 participants at risk. |
| **Alternative inactivity thresholds** | | 180-day inactivity; reactivation within 365 days | 10.8% (95% CI 10.2% to 11.4%) | | 8,077 participants at risk. |
| **Alternative inactivity thresholds** | | 180-day inactivity; reactivation within 730 days | 13.1% (95% CI 12.5% to 13.8%) | | 7,279 participants at risk. |
| **Alternative inactivity thresholds** | | 365-day inactivity; reactivation within 183 days | 3.6% (95% CI 3.3% to 4.0%) | | 8,456 participants at risk. |
| **Alternative inactivity thresholds** | | 365-day inactivity; reactivation within 365 days | 5.2% (95% CI 4.8% to 5.7%) | | 7,983 participants at risk. |
| **Alternative inactivity thresholds** | | 365-day inactivity; reactivation within 730 days | 7.2% (95% CI 6.7% to 7.8%) | | 6,896 participants at risk. |
| **Alternative inactivity thresholds** | | 730-day inactivity; reactivation within 183 days | 1.3% (95% CI 1.1% to 1.6%) | | 7,738 participants at risk. |
| **Alternative inactivity thresholds** | | 730-day inactivity; reactivation within 365 days | 2.2% (95% CI 1.9% to 2.5%) | | 6,972 participants at risk. |
| **Alternative inactivity thresholds** | | 730-day inactivity; reactivation within 730 days | 3.5% (95% CI 3.1% to 4.0%) | | 5,207 participants at risk. |
| **Post-reactivation contribution** | | At least 2 additional activity days within 365 days | 203/666 (30.5%) | | Event-level denominator restricted to 365 days of subsequent follow-up opportunity. |
| **Score-related selective observation** | CFI | Prior score effect on next-assessment observation | Adjusted OR per 1-SD worse prior score 0.952 (95% CI 0.929 to 0.976); | 168,263 risk intervals; 9,611 participants | Denominator logistic observation model; participant-clustered sandwich confidence interval. |
| **Score-related selective observation** | Cogstate | Prior score effect on next-assessment observation | Adjusted OR per 1-SD worse prior score 0.845 (95% CI 0.819 to 0.872); | 102,479 risk intervals; 5,894 participants | Denominator logistic observation model; participant-clustered sandwich confidence interval. |
| **Time to first >=365-day inactivity gap** | | CFI: without a prolonged-inactivity gap at year 1 | 49.9% (95% CI 48.9% to 50.8%) | | Number at risk: 10,641. |
| **Time to first >=365-day inactivity gap** | | CFI: without a prolonged-inactivity gap at year 3 | 20.3% (95% CI 19.5% to 21.1%) | | Number at risk: 1,998. |
| **Time to first >=365-day inactivity gap** | | CFI: without a prolonged-inactivity gap at year 5 | 13.7% (95% CI 13.0% to 14.4%) | | Number at risk: 672. |
| **Time to first >=365-day inactivity gap** | | CFI: median time to first gap | 1.00 years | | Kaplan-Meier median. |
| **Time to first >=365-day inactivity gap** | | Cogstate: without a prolonged-inactivity gap at | 59.9% (95% CI 58.7% to 61.1%) | | Number at risk: 6,399. |
| **Time to first >=365-day inactivity gap** | | Cogstate: without a prolonged-inactivity gap at | 29.6% (95% CI 28.4% to 30.7%) | | Number at risk: 1,745. |
| **Time to first >=365-day inactivity gap** | | Cogstate: without a prolonged-inactivity gap at | 20.8% (95% CI 19.7% to 21.9%) | | Number at risk: 608. |
| **Time to first >=365-day inactivity gap** | | Cogstate: median time to first gap | 1.37 years | | Kaplan-Meier median. |
| **Recent-activity history and predicted next-observation probability** | CFI | Recency category <90 days | 22.8% | 168,263 modeled rolling availability intervals | Recency set to 82.0 days; predicted probabilities averaged over the observation-model covariate distribution. Descriptive association, not a causal effect. |
| **Recent-activity history and predicted next-observation probability** | CFI | Recency category 90-179 days | 22.7% | 168,263 modeled rolling availability intervals | Recency set to 90.0 days; predicted probabilities averaged over the observation-model covariate distribution. Descriptive association, not a causal effect. |
| **Recent-activity history and predicted next-observation probability** | CFI | Recency category 180-364 days | 21.2% | 168,263 modeled rolling availability intervals | Recency set to 270.0 days; predicted probabilities averaged over the observation-model covariate distribution. Descriptive association, not a causal effect. |
| **Recent-activity history and predicted next-observation probability** | CFI | Recency category 365-729 days | 18.8% | 168,263 modeled rolling availability intervals | Recency set to 540.0 days; predicted probabilities averaged over the observation-model covariate distribution. Descriptive association, not a causal effect. |
| **Recent-activity history and predicted next-observation probability** | CFI | Recency category >=730 days | 13.1% | 168,263 modeled rolling availability intervals | Recency set to 1175.0 days; predicted probabilities averaged over the observation-model covariate distribution. Descriptive association, not a causal effect. |
| **Recent-activity history and predicted next-observation probability** | Cogstate composite | Recency category <90 days | 34.3% | 102,479 modeled rolling availability intervals | Recency set to 35.0 days; predicted probabilities averaged over the observation-model covariate distribution. Descriptive association, not a causal effect. |
| **Recent-activity history and predicted next-observation probability** | Cogstate composite | Recency category 90-179 days | 32.5% | 102,479 modeled rolling availability intervals | Recency set to 90.0 days; predicted probabilities averaged over the observation-model covariate distribution. Descriptive association, not a causal effect. |
| **Recent-activity history and predicted next-observation probability** | Cogstate composite | Recency category 180-364 days | 26.5% | 102,479 modeled rolling availability intervals | Recency set to 270.0 days; predicted probabilities averaged over the observation-model covariate distribution. Descriptive association, not a causal effect. |
| **Recent-activity history and predicted next-observation probability** | Cogstate composite | Recency category 365-729 days | 17.2% | 102,479 modeled rolling availability intervals | Recency set to 540.0 days; predicted probabilities averaged over the observation-model covariate distribution. Descriptive association, not a causal effect. |
| **Recent-activity history and predicted next-observation probability** | Cogstate composite | Recency category >=730 days | 2.6% | 102,479 modeled rolling availability intervals | Recency set to 1170.0 days; predicted probabilities averaged over the observation-model covariate distribution. Descriptive association, not a causal effect. |
| **Temporal replication of engagement** | | 2022-2023 registration cohort: At least one valid assessment | 97.9% | 5,212 eligible participants | Both registration cohorts were evaluated within a common 24-month observation window. |
| **Temporal replication of engagement** | | 2022-2023 registration cohort: At least one valid CFI assessment | 97.9% | 5,212 eligible participants | Both registration cohorts were evaluated within a common 24-month observation window. |
| **Temporal replication of engagement** | | 2022-2023 registration cohort: Repeated CFI assessment | 51.9% | 5,212 eligible participants | Both registration cohorts were evaluated within a common 24-month observation window. |
| **Temporal replication of engagement** | | 2022-2023 registration cohort: At least one valid Cogstate assessment | 57.8% | 5,212 eligible participants | Both registration cohorts were evaluated within a common 24-month observation window. |
| **Temporal replication of engagement** | | 2022-2023 registration cohort: Repeated Cogstate assessment | 36.0% | 5,212 eligible participants | Both registration cohorts were evaluated within a common 24-month observation window. |
| **Temporal replication of engagement** | | 2022-2023 registration cohort: Contribution at or beyond 12 months | 32.3% | 5,212 eligible participants | Both registration cohorts were evaluated within a common 24-month observation window. |
| **Temporal replication of engagement** | | 2019-2021 registration cohort: At least one valid assessment | 98.1% | 5,133 eligible participants | Both registration cohorts were evaluated within a common 24-month observation window. |
| **Temporal replication of engagement** | | 2019-2021 registration cohort: At least one valid CFI assessment | 98.1% | 5,133 eligible participants | Both registration cohorts were evaluated within a common 24-month observation window. |
| **Temporal replication of engagement** | | 2019-2021 registration cohort: Repeated CFI assessment | 51.8% | 5,133 eligible participants | Both registration cohorts were evaluated within a common 24-month observation window. |
| **Temporal replication of engagement** | | 2019-2021 registration cohort: At least one valid Cogstate assessment | 60.4% | 5,133 eligible participants | Both registration cohorts were evaluated within a common 24-month observation window. |
| **Temporal replication of engagement** | | 2019-2021 registration cohort: Repeated Cogstate assessment | 38.1% | 5,133 eligible participants | Both registration cohorts were evaluated within a common 24-month observation window. |
| **Temporal replication of engagement** | | 2019-2021 registration cohort: Contribution at or beyond 12 months | 34.9% | 5,133 eligible participants | Both registration cohorts were evaluated within a common 24-month observation window. |

**Legend:**

Long-term contribution requires sufficient follow-up opportunity. Activity states and reactivation probabilities have different denominators/risk sets; Kaplan-Meier estimates account for censoring. Prior-score and recency associations are noncausal.

**Abbreviations:**

CFI, Cognitive Function Instrument; CI, confidence interval; OR, odds ratio; SD, standard deviation.

**Supplementary Table S3. Comparable 0-36-month retest/time model grid**

| Outcome | Elapsed-time function | Retest representation | Estimator | Time association, favorable SD/year (95% CI) | Participants | Assessment records | Delta AIC within outcome/time/estimator | Free CV RMSE (native units; marginal only) | Free CV excess (%; marginal only) | CV RMSE change with reference-constrained time coefficient (%; linear marginal only) |
|---|---|---|---|---|---|---|---|---|---|---|
| CFI | Linear | Assessment number 1/2/3/4+ | Working-independence marginal | 0.080 (0.061 to 0.099) | 9611 | 36626 | 111.86 | 1.8145 | 0.1 | 0.373 |
| CFI | Linear | Assessment number 1/2/3/4+ | LMM ML | 0.078 (0.064 to 0.092) | 9611 | 36626 | 76.12 | | | |
| CFI | Linear | Assessment number 1/2/3/4+ | Participant fixed effects | 0.081 (0.068 to 0.093) | 5052 | 32067 | | | | |
| CFI | Linear | Post-first + linear log N | Working-independence marginal | -0.117 (-0.172 to -0.062) | 9611 | 36626 | 24.47 | 1.8127 | 0 | -0.02 |
| CFI | Linear | Post-first + linear log N | LMM ML | -0.041 (-0.068 to -0.015) | 9611 | 36626 | 8.82 | | | |
| CFI | Linear | Post-first + linear log N | Participant fixed effects | -0.039 (-0.068 to -0.011) | 5052 | 32067 | | | | |
| CFI | Linear | Expanded assessment-number categories | Working-independence marginal | -0.129 (-0.199 to -0.058) | 9611 | 36626 | 40.05 | 1.8138 | 0.057 | 0.006 |
| CFI | Linear | Expanded assessment-number categories | LMM ML | -0.013 (-0.044 to 0.019) | 9611 | 36626 | 31.94 | | | |
| CFI | Linear | Expanded assessment-number categories | Participant fixed effects | -0.018 (-0.052 to 0.015) | 5052 | 32067 | | | | |
| CFI | Linear | Exact assessment number through 10 | Working-independence marginal | -0.216 (-0.312 to -0.119) | 9611 | 36626 | 15.03 | 1.8148 | 0.113 | -0.005 |
| CFI | Linear | Exact assessment number through 10 | LMM ML | -0.102 (-0.148 to -0.057) | 9611 | 36626 | 7.84 | | | |
| CFI | Linear | Exact assessment number through 10 | Participant fixed effects | -0.086 (-0.138 to -0.035) | 5052 | 32067 | | | | |
| CFI | Linear | Post-first + spline log N | Working-independence marginal | -0.227 (-0.327 to -0.127) | 9611 | 36626 | 0 | 1.815 | 0.125 | -0.005 |
| CFI | Linear | Post-first + spline log N | LMM ML | -0.115 (-0.163 to -0.067) | 9611 | 36626 | 0 | | | |
| CFI | Linear | Post-first + spline log N | Participant fixed effects | -0.094 (-0.149 to -0.039) | 5052 | 32067 | | | | |
| CFI | Spline; annualized 0-36-month contrast | Assessment number 1/2/3/4+ | Working-independence marginal | 0.132 (0.030 to 0.234) | 9611 | 36626 | 112.06 | 1.8141 | 0.007 | |
| CFI | Spline; annualized 0-36-month contrast | Assessment number 1/2/3/4+ | LMM ML | 0.188 (0.136 to 0.239) | 9611 | 36626 | 66.35 | | | |
| CFI | Spline; annualized 0-36-month contrast | Assessment number 1/2/3/4+ | Participant fixed effects | 0.189 (0.138 to 0.240) | 5052 | 32067 | | | | |
| CFI | Spline; annualized 0-36-month contrast | Post-first + linear log N | Working-independence marginal | -0.240 (-0.372 to -0.109) | 9611 | 36626 | 10.2 | 1.8152 | 0.065 | |
| CFI | Spline; annualized 0-36-month contrast | Post-first + linear log N | LMM ML | -0.073 (-0.136 to -0.011) | 9611 | 36626 | 19.74 | | | |
| CFI | Spline; annualized 0-36-month contrast | Post-first + linear log N | Participant fixed effects | -0.066 (-0.137 to 0.006) | 5052 | 32067 | | | | |
| CFI | Spline; annualized 0-36-month contrast | Expanded assessment-number categories | Working-independence marginal | -0.053 (-0.176 to 0.070) | 9611 | 36626 | 34.37 | 1.814 | 0 | |
| CFI | Spline; annualized 0-36-month contrast | Expanded assessment-number categories | LMM ML | 0.088 (0.027 to 0.148) | 9611 | 36626 | 21.03 | | | |
| CFI | Spline; annualized 0-36-month contrast | Expanded assessment-number categories | Participant fixed effects | 0.078 (0.014 to 0.142) | 5052 | 32067 | | | | |
| CFI | Spline; annualized 0-36-month contrast | Exact assessment number through 10 | Working-independence marginal | -0.140 (-0.281 to 0.001) | 9611 | 36626 | 13.93 | 1.8148 | 0.042 | |
| CFI | Spline; annualized 0-36-month contrast | Exact assessment number through 10 | LMM ML | 0.013 (-0.056 to 0.082) | 9611 | 36626 | 0 | | | |
| CFI | Spline; annualized 0-36-month contrast | Exact assessment number through 10 | Participant fixed effects | 0.020 (-0.054 to 0.094) | 5052 | 32067 | | | | |
| CFI | Spline; annualized 0-36-month contrast | Post-first + spline log N | Working-independence marginal | -0.166 (-0.303 to -0.029) | 9611 | 36626 | 0 | 1.8149 | 0.05 | |
| CFI | Spline; annualized 0-36-month contrast | Post-first + spline log N | LMM ML | -0.026 (-0.094 to 0.042) | 9611 | 36626 | 0.95 | | | |
| CFI | Spline; annualized 0-36-month contrast | Post-first + spline log N | Participant fixed effects | -0.016 (-0.089 to 0.058) | 5052 | 32067 | | | | |
| Cogstate composite | Linear | Assessment number 1/2/3/4+ | Working-independence marginal | 0.265 (0.245 to 0.285) | 5898 | 27775 | 91.5 | 0.525 | 0.375 | 1.121 |
| Cogstate composite | Linear | Assessment number 1/2/3/4+ | LMM ML | 0.248 (0.231 to 0.265) | 5898 | 27775 | 98.95 | | | |
| Cogstate composite | Linear | Assessment number 1/2/3/4+ | Participant fixed effects | 0.252 (0.235 to 0.269) | 3646 | 25523 | | | | |
| Cogstate composite | Linear | Post-first + linear log N | Working-independence marginal | 0.033 (-0.023 to 0.090) | 5898 | 27775 | 9.72 | 0.5231 | 0.014 | 0.005 |
| Cogstate composite | Linear | Post-first + linear log N | LMM ML | 0.049 (0.012 to 0.086) | 5898 | 27775 | 12.41 | | | |
| Cogstate composite | Linear | Post-first + linear log N | Participant fixed effects | 0.052 (0.012 to 0.091) | 3646 | 25523 | | | | |
| Cogstate composite | Linear | Expanded assessment-number categories | Working-independence marginal | 0.057 (-0.014 to 0.128) | 5898 | 27775 | 34.78 | 0.5235 | 0.088 | 0.071 |
| Cogstate composite | Linear | Expanded assessment-number categories | LMM ML | 0.092 (0.049 to 0.136) | 5898 | 27775 | 43.65 | | | |
| Cogstate composite | Linear | Expanded assessment-number categories | Participant fixed effects | 0.079 (0.032 to 0.126) | 3646 | 25523 | | | | |
| Cogstate composite | Linear | Exact assessment number through 10 | Working-independence marginal | -0.033 (-0.133 to 0.066) | 5898 | 27775 | 16.71 | 0.5231 | 0.02 | 0.04 |
| Cogstate composite | Linear | Exact assessment number through 10 | LMM ML | -0.032 (-0.093 to 0.030) | 5898 | 27775 | 16.41 | | | |
| Cogstate composite | Linear | Exact assessment number through 10 | Participant fixed effects | -0.043 (-0.113 to 0.026) | 3646 | 25523 | | | | |
| Cogstate composite | Linear | Post-first + spline log N | Working-independence marginal | -0.049 (-0.153 to 0.055) | 5898 | 27775 | 0 | 0.523 | 0 | 0.027 |
| Cogstate composite | Linear | Post-first + spline log N | LMM ML | -0.060 (-0.125 to 0.004) | 5898 | 27775 | 0 | | | |
| Cogstate composite | Linear | Post-first + spline log N | Participant fixed effects | -0.071 (-0.144 to 0.003) | 3646 | 25523 | | | | |
| Cogstate composite | Spline; annualized 0-36-month contrast | Assessment number 1/2/3/4+ | Working-independence marginal | 0.224 (0.136 to 0.313) | 5898 | 27775 | 88.68 | 0.525 | 0.377 | |
| Cogstate composite | Spline; annualized 0-36-month contrast | Assessment number 1/2/3/4+ | LMM ML | 0.219 (0.158 to 0.280) | 5898 | 27775 | 92.84 | | | |
| Cogstate composite | Spline; annualized 0-36-month contrast | Assessment number 1/2/3/4+ | Participant fixed effects | 0.269 (0.207 to 0.332) | 3646 | 25523 | | | | |
| Cogstate composite | Spline; annualized 0-36-month contrast | Post-first + linear log N | Working-independence marginal | -0.078 (-0.213 to 0.057) | 5898 | 27775 | 1.95 | 0.523 | 0 | |
| Cogstate composite | Spline; annualized 0-36-month contrast | Post-first + linear log N | LMM ML | -0.099 (-0.183 to -0.014) | 5898 | 27775 | 0 | | | |
| Cogstate composite | Spline; annualized 0-36-month contrast | Post-first + linear log N | Participant fixed effects | -0.118 (-0.217 to -0.019) | 3646 | 25523 | | | | |
| Cogstate composite | Spline; annualized 0-36-month contrast | Expanded assessment-number categories | Working-independence marginal | 0.012 (-0.108 to 0.131) | 5898 | 27775 | 34.64 | 0.5235 | 0.09 | |
| Cogstate composite | Spline; annualized 0-36-month contrast | Expanded assessment-number categories | LMM ML | 0.036 (-0.042 to 0.113) | 5898 | 27775 | 42.79 | | | |
| Cogstate composite | Spline; annualized 0-36-month contrast | Expanded assessment-number categories | Participant fixed effects | 0.055 (-0.030 to 0.140) | 3646 | 25523 | | | | |
| Cogstate composite | Spline; annualized 0-36-month contrast | Exact assessment number through 10 | Working-independence marginal | -0.086 (-0.223 to 0.050) | 5898 | 27775 | 15.24 | 0.5231 | 0.023 | |
| Cogstate composite | Spline; annualized 0-36-month contrast | Exact assessment number through 10 | LMM ML | -0.081 (-0.169 to 0.008) | 5898 | 27775 | 16.75 | | | |
| Cogstate composite | Spline; annualized 0-36-month contrast | Exact assessment number through 10 | Participant fixed effects | -0.076 (-0.175 to 0.024) | 3646 | 25523 | | | | |
| Cogstate composite | Spline; annualized 0-36-month contrast | Post-first + spline log N | Working-independence marginal | -0.076 (-0.211 to 0.060) | 5898 | 27775 | 0 | 0.523 | 0.007 | |
| Cogstate composite | Spline; annualized 0-36-month contrast | Post-first + spline log N | LMM ML | -0.096 (-0.183 to -0.009) | 5898 | 27775 | 1.3 | | | |
| Cogstate composite | Spline; annualized 0-36-month contrast | Post-first + spline log N | Participant fixed effects | -0.094 (-0.194 to 0.006) | 3646 | 25523 | | | | |

**Legend:**

Five retest representations, two elapsed-time functions, and three estimator classes were evaluated for each outcome. Time coefficients are in favorable baseline SD/year; spline-time contrasts are annualized over months 0-36. Participant fixed-effects models exclude participants with only one observed assessment. Delta AIC comparisons are restricted to the same outcome, estimator, and elapsed-time function (fixed-effects AIC was not evaluated). Participant-clustered cross-validation was performed for marginal models only; RMSE is expressed in the original outcome units. Blank cross-validation entries indicate analyses that were not performed. For linear-time marginal models, the reference-constrained cross-validation is a prediction-sensitivity diagnostic, not validation of the first-exposure reference as a biological time slope.

**Abbreviations:**

AIC, Akaike information criterion; CFI, Cognitive Function Instrument; CI, confidence interval; CV, cross-validation; FE, fixed effects; LMM, linear mixed model; RMSE, root mean square error; SD, standard deviation.

### Supplementary Table S4, Part A. Primary matching by assessment number

| Outcome | Assessment number | Median elapsed (years) | Eligible returnees | Matched pairs | Matched (%) | Returnee change, favorable SD (95% CI) | First-exposure change, favorable SD (95% CI) | Difference of changes, favorable SD (95% CI) |
|---|---|---|---|---|---|---|---|---|
| CFI | 2 | 0.26 | 5554 | 4262 | 76.7 | 0.105 (0.083 to 0.126) | -0.043 (-0.084 to -0.002) | 0.147 (0.101 to 0.194) |
| CFI | 3 | 0.52 | 4149 | 3626 | 87.4 | 0.196 (0.172 to 0.220) | -0.035 (-0.080 to 0.011) | 0.230 (0.179 to 0.282) |
| CFI | 4 | 0.79 | 3446 | 2967 | 86.1 | 0.233 (0.206 to 0.260) | -0.047 (-0.098 to 0.003) | 0.281 (0.223 to 0.338) |
| CFI | 5 | 1.05 | 2999 | 2584 | 86.2 | 0.274 (0.245 to 0.303) | -0.039 (-0.094 to 0.016) | 0.313 (0.251 to 0.375) |
| CFI | 6 | 1.31 | 2648 | 2282 | 86.2 | 0.320 (0.289 to 0.351) | -0.051 (-0.108 to 0.007) | 0.371 (0.306 to 0.436) |
| CFI | 7 | 1.57 | 2363 | 1989 | 84.2 | 0.326 (0.291 to 0.360) | -0.119 (-0.182 to -0.056) | 0.444 (0.373 to 0.515) |
| CFI | 8 | 1.83 | 2145 | 1712 | 79.8 | 0.366 (0.329 to 0.402) | -0.067 (-0.134 to 0.001) | 0.432 (0.357 to 0.508) |
| CFI | 9 | 2.09 | 1923 | 1537 | 79.9 | 0.375 (0.335 to 0.415) | -0.086 (-0.158 to -0.013) | 0.461 (0.378 to 0.543) |
| CFI | 10 | 2.35 | 1722 | 1363 | 79.2 | 0.395 (0.353 to 0.436) | -0.055 (-0.133 to 0.022) | 0.450 (0.362 to 0.538) |
| CFI | 11 | 2.6 | 1480 | 1203 | 81.3 | 0.419 (0.374 to 0.465) | -0.044 (-0.126 to 0.039) | 0.463 (0.369 to 0.557) |
| CFI | 12 | 2.83 | 1112 | 931 | 83.7 | 0.433 (0.381 to 0.484) | -0.058 (-0.149 to 0.033) | 0.491 (0.385 to 0.596) |
| Cogstate composite | 2 | 0.27 | 3974 | 2202 | 55.4 | 0.220 (0.183 to 0.257) | -0.040 (-0.099 to 0.019) | 0.260 (0.190 to 0.329) |
| Cogstate composite | 3 | 0.53 | 3203 | 2543 | 79.4 | 0.388 (0.354 to 0.421) | -0.021 (-0.075 to 0.033) | 0.409 (0.347 to 0.471) |
| Cogstate composite | 4 | 0.79 | 2767 | 2148 | 77.6 | 0.520 (0.483 to 0.558) | 0.046 (-0.012 to 0.105) | 0.474 (0.404 to 0.543) |
| Cogstate composite | 5 | 1.06 | 2460 | 1845 | 75 | 0.626 (0.584 to 0.668) | 0.073 (0.009 to 0.136) | 0.553 (0.477 to 0.630) |
| Cogstate composite | 6 | 1.32 | 2206 | 1663 | 75.4 | 0.713 (0.670 to 0.756) | 0.085 (0.019 to 0.150) | 0.628 (0.550 to 0.706) |
| Cogstate composite | 7 | 1.59 | 2011 | 1512 | 75.2 | 0.778 (0.731 to 0.824) | 0.020 (-0.050 to 0.091) | 0.757 (0.672 to 0.843) |
| Cogstate composite | 8 | 1.85 | 1849 | 1324 | 71.6 | 0.857 (0.808 to 0.907) | 0.041 (-0.035 to 0.116) | 0.817 (0.726 to 0.908) |
| Cogstate composite | 9 | 2.11 | 1672 | 1167 | 69.8 | 0.902 (0.848 to 0.956) | 0.054 (-0.027 to 0.136) | 0.848 (0.750 to 0.945) |
| Cogstate composite | 10 | 2.36 | 1493 | 991 | 66.4 | 0.990 (0.929 to 1.050) | 0.044 (-0.044 to 0.133) | 0.945 (0.836 to 1.055) |
| Cogstate composite | 11 | 2.62 | 1285 | 843 | 65.6 | 1.037 (0.970 to 1.104) | 0.099 (0.006 to 0.192) | 0.938 (0.825 to 1.051) |
| Cogstate composite | 12 | 2.84 | 926 | 628 | 67.8 | 1.140 (1.063 to 1.217) | 0.080 (-0.029 to 0.189) | 1.060 (0.925 to 1.196) |

**Legend:**

Primary 1:1 contemporaneous matching at assessment number 2-12 within 36 months. Changes compare returnees with separately observed first J-TRC exposures at matched calendar dates. Favorable standardized direction is positive. Assessment-specific CIs are not rematching-bootstrap CIs. The matched comparator is neither a same-person counterfactual nor a pure aging slope.

**Abbreviations:**

CFI, Cognitive Function Instrument; CI, confidence interval; SD, baseline standard deviation.

### Supplementary Table S4, Part B. Reference slopes and robustness

| Section | Outcome | Analysis | Estimate / statistic | Units | Participant / variable count | Assessment observations | Valid / requested bootstrap | Notes |
|---|---|---|---|---|---|---|---|---|
| **Primary first-exposure reference** | CFI | Reference slope (participant-equal; rematching bootstrap CI) | -0.033 (-0.057 to -0.007) | Favorable baseline SD/year | 4835 | 24456 | 500/500 | First recorded J-TRC assessment; prior testing outside this registry is unknown. Not |
| **Primary first-exposure reference** | Cogstate composite | Reference slope (participant-equal; rematching bootstrap CI) | 0.025 (-0.005 to 0.054) | Favorable baseline SD/year | 3250 | 16866 | 500/500 | First recorded J-TRC assessment; prior testing outside this registry is unknown. Not |
| **First-assessment timing sensitivity** | CFI | First exposure within 90 days of registration (rematching bootstrap CI) | -0.038 (-0.058 to -0.009) | Favorable baseline SD/year | 4785 | 24200 | 500/500 | Comparator first assessments restricted to within 90 days of registration. |
| **First-assessment timing sensitivity** | Cogstate composite | First exposure within 90 days of registration (rematching bootstrap CI) | 0.021 (-0.004 to 0.058) | Favorable baseline SD/year | 2997 | 15171 | 500/500 | Comparator first assessments restricted to within 90 days of registration. |
| **Matching-calendar-window sensitivity** | CFI | Reference slope, calendar +/-90 days (cluster-robust CI) | -0.024 (-0.045 to -0.003) | Favorable baseline SD/year | 4704 | 21662 | Not a bootstrap CI | Alternative matching window; do not equate robust and rematching-bootstrap CI |
| **Matching-calendar-window sensitivity** | Cogstate composite | Reference slope, calendar +/-90 days (cluster-robust CI) | 0.020 (-0.004 to 0.045) | Favorable baseline SD/year | 3145 | 14602 | Not a bootstrap CI | Alternative matching window; do not equate robust and rematching-bootstrap CI |
| **Measured later-entrant composition (descriptive)** | CFI | Maximum absolute early/late standardized difference | 0.134 | Absolute standardized mean difference | 13 | | Not applicable | Variable count, not participant count; unmeasured differences between enrollment |
| **Measured later-entrant composition (descriptive)** | Cogstate composite | Maximum absolute early/late standardized difference | 0.137 | Absolute standardized mean difference | 13 | | Not applicable | Variable count, not participant count; unmeasured differences between enrollment |

**Legend:**

The primary reference uses participant-level bootstrap with repeated matching. When included, the sensitivity restricting first recorded assessments to within 90 days of registration uses the same inference approach; valid and requested replicate counts are reported. The narrower calendar-matching-window analysis uses participant-clustered robust confidence intervals instead. The maximum standardized difference between earlier and later entrants describes measured covariates and does not exclude unmeasured calendar-period differences. The reference is not a within-participant biological aging trajectory.

**Abbreviations:**

CFI, Cognitive Function Instrument; CI, confidence interval; SD, baseline standard deviation; SMD, standardized mean difference.

### Supplementary Table S4, Part C. IPOW and observation sensitivity

| Section | Outcome | Analysis | Estimate or result | Sample | Additional information |
|---|---|---|---|---|---|
| **Observation-panel IPOW diagnostics** | CFI | Primary 1st–99th percentile truncation | ESS 6371.6 (13.3% of observed weighted assessments) | 186,081 rolling availability intervals; 47,750 observed weighted assessments | 49.9% of modeled rolling availability intervals had denominator probability <0.01; raw maximum weight 1540.8. |
| **Observation-panel IPOW diagnostics** | Cogstate composite | Primary 1st–99th percentile truncation | ESS 9940.3 (26.8% of observed weighted assessments) | 111,299 rolling availability intervals; 37,135 observed weighted assessments | 43.4% of modeled rolling availability intervals had denominator probability <0.01; raw maximum weight 2408.2. |
| **Flexible observation-model functional-form sensitivity** | CFI | Natural-spline elapsed time + natural-spline log recency | IPOW -0.083 (-0.154 to -0.013); ESS 6396.1 (13.7%) | 168,263 modeled noninitial rolling availability intervals; 46,716 weighted outcome records | 52.9% of modeled noninitial intervals had denominator probability <0.01. The stabilized numerator from the primary model was held fixed while the denominator functional form was varied. |
| **Flexible observation-model functional-form sensitivity** | Cogstate composite | Natural-spline elapsed time + natural-spline log recency | IPOW 0.096 (0.082 to 0.109); ESS 8478.3 (23.2%) | 102,479 modeled noninitial rolling availability intervals; 36,614 weighted outcome records | 44.9% of modeled noninitial intervals had denominator probability <0.01. The stabilized numerator from the primary model was held fixed while the denominator functional form was varied. |
| **Complete-case outcome-model weight sensitivity** | CFI | p01_p99_primary | Elapsed-time association -0.084 (-0.154 to -0.014); ESS 6054.1 (13.0%) | 9,611 participants; 46,716 complete-case weighted outcome records | 2.0% of observed interval-specific weights truncated. Weights are not cumulatively multiplied across prior intervals; confidence intervals condition on the estimated weights. |
| **Complete-case outcome-model weight sensitivity** | CFI | p025_p975 | Elapsed-time association -0.063 (-0.099 to -0.027); ESS 28295.1 (60.6%) | 9,611 participants; 46,716 complete-case weighted outcome records | 5.0% of observed interval-specific weights truncated. Weights are not cumulatively multiplied across prior intervals; confidence intervals condition on the estimated weights. |
| **Complete-case outcome-model weight sensitivity** | CFI | p05_p95 | Elapsed-time association -0.059 (-0.092 to -0.026); ESS 32714.8 (70.0%) | 9,611 participants; 46,716 complete-case weighted outcome records | 8.2% of observed interval-specific weights truncated. Weights are not cumulatively multiplied across prior intervals; confidence intervals condition on the estimated weights. |
| **Complete-case outcome-model weight sensitivity** | Cogstate composite | p01_p99_primary | Elapsed-time association 0.098 (0.085 to 0.111); ESS 9615.6 (26.3%) | 5,898 participants; 36,614 complete-case weighted outcome records | 2.0% of observed interval-specific weights truncated. Weights are not cumulatively multiplied across prior intervals; confidence intervals condition on the estimated weights. |
| **Complete-case outcome-model weight sensitivity** | Cogstate composite | p025_p975 | Elapsed-time association 0.098 (0.089 to 0.108); ESS 24470.0 (66.8%) | 5,898 participants; 36,614 complete-case weighted outcome records | 5.0% of observed interval-specific weights truncated. Weights are not cumulatively multiplied across prior intervals; confidence intervals condition on the estimated weights. |
| **Complete-case outcome-model weight sensitivity** | Cogstate composite | p05_p95 | Elapsed-time association 0.100 (0.091 to 0.109); ESS 28191.0 (77.0%) | 5,898 participants; 36,614 complete-case weighted outcome records | 8.5% of observed interval-specific weights truncated. Weights are not cumulatively multiplied across prior intervals; confidence intervals condition on the estimated weights. |
| **Risk-bin-width sensitivity of reference IPOW analysis** | CFI | 60-day risk-bin width | Unweighted -0.066 (-0.097 to -0.035); IPOW -0.091 (-0.170 to -0.013) | 9,611 participants; 46,716 weighted intervals | ESS fraction 12.3%; 59.1% of modeled intervals had denominator probability <0.01. Risk intervals began at rolling availability dates; complete censored intervals were included and terminal censored partial intervals were excluded. |
| **Risk-bin-width sensitivity of reference IPOW analysis** | CFI | 90-day risk-bin width | Unweighted -0.066 (-0.097 to -0.035); IPOW -0.084 (-0.154 to -0.014) | 9,611 participants; 46,716 weighted intervals | ESS fraction 13.3%; 49.9% of modeled intervals had denominator probability <0.01. Risk intervals began at rolling availability dates; complete censored intervals were included and terminal censored partial intervals were excluded. |
| **Risk-bin-width sensitivity of reference IPOW analysis** | CFI | 120-day risk-bin width | Unweighted -0.066 (-0.097 to -0.035); IPOW -0.090 (-0.156 to -0.024) | 9,611 participants; 46,716 weighted intervals | ESS fraction 13.2%; 43.4% of modeled intervals had denominator probability <0.01. Risk intervals began at rolling availability dates; complete censored intervals were included and terminal censored partial intervals were excluded. |
| **Risk-bin-width sensitivity of reference IPOW analysis** | Cogstate composite | 60-day risk-bin width | Unweighted 0.103 (0.095 to 0.111); IPOW 0.097 (0.082 to 0.112) | 5,898 participants; 36,614 weighted intervals | ESS fraction 21.9%; 52.1% of modeled intervals had denominator probability <0.01. Risk intervals began at rolling availability dates; complete censored intervals were included and terminal censored partial intervals were excluded. |
| **Risk-bin-width sensitivity of reference IPOW analysis** | Cogstate composite | 90-day risk-bin width | Unweighted 0.103 (0.095 to 0.111); IPOW 0.098 (0.085 to 0.111) | 5,898 participants; 36,614 weighted intervals | ESS fraction 26.8%; 43.4% of modeled intervals had denominator probability <0.01. Risk intervals began at rolling availability dates; complete censored intervals were included and terminal censored partial intervals were excluded. |
| **Risk-bin-width sensitivity of reference IPOW analysis** | Cogstate composite | 120-day risk-bin width | Unweighted 0.103 (0.095 to 0.111); IPOW 0.101 (0.089 to 0.113) | 5,898 participants; 36,614 weighted intervals | ESS fraction 26.7%; 36.1% of modeled intervals had denominator probability <0.01. Risk intervals began at rolling availability dates; complete censored intervals were included and terminal censored partial intervals were excluded. |
| **Common 36-month retention-pattern sensitivity** | CFI | Categorical assessment number (1/2/3/4+) | -0.018 (-0.210 to 0.173) | 8,831 participants; 34,242 90-day analysis records | Retention group defined by assessment count accrued within the first 36 months among participants with at least 36 months of follow-up opportunity. |
| **Common 36-month retention-pattern sensitivity** | CFI | Post-first + linear log assessment number | 0.212 (0.013 to 0.410) | 8,831 participants; 34,242 90-day analysis records | Retention group defined by assessment count accrued within the first 36 months among participants with at least 36 months of follow-up opportunity. |
| **Common 36-month retention-pattern sensitivity** | CFI | Post-first + spline of log assessment number | 0.343 (0.126 to 0.561) | 8,831 participants; 34,242 90-day analysis records | Retention group defined by assessment count accrued within the first 36 months among participants with at least 36 months of follow-up opportunity. |
| **Common 36-month retention-pattern sensitivity** | Cogstate composite | Categorical assessment number (1/2/3/4+) | 0.113 (0.077 to 0.150) | 5,410 participants; 25,765 90-day analysis records | Retention group defined by assessment count accrued within the first 36 months among participants with at least 36 months of follow-up opportunity. |
| **Common 36-month retention-pattern sensitivity** | Cogstate composite | Post-first + linear log assessment number | 0.040 (-0.000 to 0.081) | 5,410 participants; 25,765 90-day analysis records | Retention group defined by assessment count accrued within the first 36 months among participants with at least 36 months of follow-up opportunity. |
| **Common 36-month retention-pattern sensitivity** | Cogstate composite | Post-first + spline of log assessment number | 0.018 (-0.028 to 0.063) | 5,410 participants; 25,765 90-day analysis records | Retention group defined by assessment count accrued within the first 36 months among participants with at least 36 months of follow-up opportunity. |
| **Joint sensitivity to prior-assessment modeling and selective follow-up** | CFI | Categorical assessment number (1/2/3/4+) | Unweighted -0.066 (-0.097 to -0.035); IPOW -0.084 (-0.154 to -0.014); retention-pattern | Representative model for prior assessment history | Primary unweighted, IPOW, and retention-pattern estimates. |
| **Joint sensitivity to prior-assessment modeling and selective follow-up** | CFI | Post-first + linear log assessment number | Unweighted 0.242 (0.175 to 0.310); IPOW 0.107 (-0.022 to 0.237); retention-pattern | Representative model for prior assessment history | Primary unweighted, IPOW, and retention-pattern estimates. |
| **Joint sensitivity to prior-assessment modeling and selective follow-up** | CFI | Post-first + spline of log assessment number | Unweighted 0.396 (0.257 to 0.534); IPOW 0.199 (0.029 to 0.369); retention-pattern | Representative model for prior assessment history | Primary unweighted, IPOW, and retention-pattern estimates. |
| **Joint sensitivity to prior-assessment modeling and selective follow-up** | Cogstate composite | Categorical assessment number (1/2/3/4+) | Unweighted 0.103 (0.095 to 0.111); IPOW 0.098 (0.085 to 0.111); retention-pattern | Representative model for prior assessment history | Primary unweighted, IPOW, and retention-pattern estimates. |
| **Joint sensitivity to prior-assessment modeling and selective follow-up** | Cogstate composite | Post-first + linear log assessment number | Unweighted 0.006 (-0.010 to 0.023); IPOW 0.017 (-0.010 to 0.044); retention-pattern | Representative model for prior assessment history | Primary unweighted, IPOW, and retention-pattern estimates. |
| **Joint sensitivity to prior-assessment modeling and selective follow-up** | Cogstate composite | Post-first + spline of log assessment number | Unweighted -0.017 (-0.046 to 0.013); IPOW 0.013 (-0.023 to 0.048); retention-pattern | Representative model for prior assessment history | Primary unweighted, IPOW, and retention-pattern estimates. |

**Legend:**

IPOW, retention, and observation-process sensitivities use registration-anchored elapsed time where specified, together with outcome-specific rolling availability/risk intervals beginning after previous completed assessments. These operational time scales and risk sets are distinct from the first-valid-assessment-anchored 0-36-month longitudinal models in Figure 4B/Table S3. The 36-month retention eligibility definition is separate from the longitudinal model inclusion window. Outcome coefficient units are native CFI score/year (higher worse) or Cogstate z/year (higher better), not the favorable baseline SD/year units of Tables S3/S5. Effective sample size, low observation probabilities and weight truncation reflect weak positivity; weighting does not identify a uniquely corrected biological trajectory. Unweighted denotes the no-IPOW analysis, not the first-exposure reference.

**Abbreviations:**

CFI, Cognitive Function Instrument; CI, confidence interval; ESS, effective sample size; IPOW, inverse probability of observation weighting; SD, standard deviation.

### Supplementary Table S5. Time-origin and first-assessment delay sensitivity

| Outcome | Retest representation | Estimator | Participants | Assessment records | Primary B: first-assessment origin (95% CI) | Sensitivity A: registration origin (95% CI) | Sensitivity C: first origin plus delay (95% CI) | A minus B (favorable SD/year) | C minus B (favorable SD/year) |
|---|---|---|---|---|---|---|---|---|---|
| CFI | Assessment number 1/2/3/4+ | Working-independence marginal | 9611 | 36626 | 0.080 (0.061 to 0.099) | 0.067 (0.047 to 0.086) | 0.080 (0.061 to 0.099) | -0.013 | 0 |
| CFI | Assessment number 1/2/3/4+ | LMM ML | 9611 | 36626 | 0.078 (0.064 to 0.092) | 0.076 (0.063 to 0.090) | 0.078 (0.064 to 0.092) | -0.002 | 0 |
| CFI | Assessment number 1/2/3/4+ | Participant fixed effects | 5052 | 32067 | 0.081 (0.068 to 0.093) | 0.081 (0.068 to 0.093) | 0.081 (0.068 to 0.093) | 0 | 0 |
| CFI | Post-first + linear log N | Working-independence marginal | 9611 | 36626 | -0.117 (-0.172 to -0.062) | -0.124 (-0.174 to -0.074) | -0.116 (-0.171 to -0.062) | -0.007 | 0.001 |
| CFI | Post-first + linear log N | LMM ML | 9611 | 36626 | -0.041 (-0.068 to -0.015) | -0.045 (-0.071 to -0.019) | -0.041 (-0.068 to -0.015) | -0.003 | 0 |
| CFI | Post-first + linear log N | Participant fixed effects | 5052 | 32067 | -0.039 (-0.068 to -0.011) | -0.039 (-0.068 to -0.011) | -0.039 (-0.068 to -0.011) | 0 | 0 |
| CFI | Post-first + spline log N | Working-independence marginal | 9611 | 36626 | -0.227 (-0.327 to -0.127) | -0.199 (-0.277 to -0.120) | -0.226 (-0.326 to -0.126) | 0.029 | 0.001 |
| CFI | Post-first + spline log N | LMM ML | 9611 | 36626 | -0.115 (-0.163 to -0.067) | -0.118 (-0.163 to -0.072) | -0.115 (-0.163 to -0.066) | -0.003 | 0 |
| CFI | Post-first + spline log N | Participant fixed effects | 5052 | 32067 | -0.094 (-0.149 to -0.039) | -0.094 (-0.149 to -0.039) | -0.094 (-0.149 to -0.039) | 0 | 0 |
| Cogstate composite | Assessment number 1/2/3/4+ | Working-independence marginal | 5898 | 27775 | 0.265 (0.245 to 0.285) | 0.183 (0.154 to 0.212) | 0.265 (0.244 to 0.285) | -0.082 | 0 |
| Cogstate composite | Assessment number 1/2/3/4+ | LMM ML | 5898 | 27775 | 0.248 (0.231 to 0.265) | 0.235 (0.218 to 0.251) | 0.248 (0.231 to 0.265) | -0.013 | 0 |
| Cogstate composite | Assessment number 1/2/3/4+ | Participant fixed effects | 3646 | 25523 | 0.252 (0.235 to 0.269) | 0.252 (0.235 to 0.269) | 0.252 (0.235 to 0.269) | 0 | 0 |
| Cogstate composite | Post-first + linear log N | Working-independence marginal | 5898 | 27775 | 0.033 (-0.023 to 0.090) | -0.014 (-0.068 to 0.039) | 0.034 (-0.022 to 0.091) | -0.048 | 0.001 |
| Cogstate composite | Post-first + linear log N | LMM ML | 5898 | 27775 | 0.049 (0.012 to 0.086) | 0.043 (0.011 to 0.075) | 0.049 (0.012 to 0.086) | -0.006 | 0 |
| Cogstate composite | Post-first + linear log N | Participant fixed effects | 3646 | 25523 | 0.052 (0.012 to 0.091) | 0.052 (0.012 to 0.091) | 0.052 (0.012 to 0.091) | 0 | 0 |
| Cogstate composite | Post-first + spline log N | Working-independence marginal | 5898 | 27775 | -0.049 (-0.153 to 0.055) | -0.041 (-0.104 to 0.022) | -0.047 (-0.150 to 0.057) | 0.008 | 0.002 |
| Cogstate composite | Post-first + spline log N | LMM ML | 5898 | 27775 | -0.060 (-0.125 to 0.004) | -0.020 (-0.066 to 0.027) | -0.060 (-0.125 to 0.004) | 0.04 | 0 |
| Cogstate composite | Post-first + spline log N | Participant fixed effects | 3646 | 25523 | -0.071 (-0.144 to 0.003) | -0.071 (-0.144 to 0.003) | -0.071 (-0.144 to 0.003) | 0 | 0 |

**Legend:**

Side-by-side comparison of representative linear-time retest models used in Figure 4B (two outcomes x three retest representations x three estimators). All A/B/C fits use identical outcome- and estimator-specific observations within 0-36 months after first valid assessment. B is the primary time since first valid assessment; A replaces this predictor with time since registration, without a delay covariate; C is time since first valid assessment plus the participant-specific registration-to-first-assessment delay (in years). Positive coefficients mean change in the favorable direction after outcome-specific baseline standardization; this is not an estimate of biological aging. Differences are A-B and C-B in favorable SD/year. Under a linear fixed-part model, A constrains the post-first time and delay coefficients to be equal, B constrains the delay coefficient to zero, and C estimates them separately. FE excludes participants with only one complete assessment. Mixed-model random-effects structures were verified separately in aggregate QC. The complete 180-fit sensitivity grid, including spline elapsed time and two additional retest models.

**Abbreviations:**

CFI, Cognitive Function Instrument; CI, confidence interval; FE, participant fixed effects; LMM, linear mixed model; SD, standard deviation.
